# Nationwide trends in COVID-19 cases and SARS-CoV-2 wastewater concentrations in the United States

**DOI:** 10.1101/2021.09.08.21263283

**Authors:** Claire Duvallet, Fuqing Wu, Kyle A. McElroy, Maxim Imakaev, Noriko Endo, Amy Xiao, Jianbo Zhang, Róisín Floyd-O’Sullivan, Morgan M Powell, Samuel Mendola, Shane T Wilson, Francis Cruz, Tamar Melman, Chaithra Lakshmi Sathyanarayana, Scott W. Olesen, Timothy B. Erickson, Newsha Ghaeli, Peter Chai, Eric Alm, Mariana Matus

**Author notes:** These authors contributed equally (Claire Duvallet, Fuqing Wu, Kyle A. McElroy, Maxim Imakaev). corresponding author Dr. Claire Duvallet.

## Abstract

Wastewater-based epidemiology has emerged as a promising technology for population-level surveillance of COVID-19 disease. The SARS-CoV-2 virus is shed in the stool of infected individuals and aggregated in public sewers, where it can be quantified to provide information on population-level disease incidence that is unbiased by access to clinical testing. In this study, we present results from the largest nationwide wastewater monitoring system in the United States reported to date. We profile 55 locations with at least six months of sampling and highlight their wastewater data from April 2020 through May 2021. These locations represent over 12 million individuals across 19 states. Samples were collected approximately weekly by wastewater treatment utilities as part of a regular wastewater surveillance service and analyzed for SARS-CoV-2 concentrations using reverse transcription quantitative polymerase chain reaction (RT-qPCR). Concentrations of SARS-CoV-2 (copies/mL) were normalized to pepper mild mottle virus (PMMoV), a stable and persistent indicator of feces concentrations in wastewater. Here, we show that wastewater data reflects temporal and geographic trends in clinical COVID-19 cases, demonstrating that wastewater surveillance is a feasible approach for nationwide population-level monitoring of COVID-19 disease. We also provide key lessons learned from our broad-scale implementation of wastewater-based epidemiology, which can be used to inform wastewater-based epidemiology approaches for future emerging diseases. With an evolving epidemic and effective vaccines against SARS-CoV-2, wastewater-based epidemiology can serve as an important passive surveillance approach to detect changing dynamics or resurgences of the virus.

**Highlights:**

- We present results from a nationwide wastewater monitoring network in the United States, which represents one of the broadest temporal and geographic wastewater-based epidemiology datasets to-date.
- Wastewater concentrations measured within individual locations reflect temporal trends in reported COVID-19 cases in the associated communities.
- Wastewater concentrations also reflect geographic patterns in reported COVID-19 cases across states throughout the pandemic.
- Normalizing wastewater concentrations to a fecal marker virus improves the correlation between wastewater data and clinical cases across locations but not necessarily over time within individual locations.
- Implementing a nationwide wastewater monitoring system for SARS-CoV-2 is feasible, practical, and sustainable.

## Introduction

The COVID-19 pandemic has galvanized the rapid development of innovative approaches for pandemic preparedness and response. Wastewater based epidemiology (WBE) to monitor for the SARS-CoV-2 virus in wastewater has emerged as a promising technology for public health surveillance. The SARS-CoV-2 virus is excreted in human feces early in the clinical course of infection and provides a view of COVID-19 that is independent of access to testing, making it an ideal candidate for WBE. WBE has previously been demonstrated in infectious disease monitoring, providing early warnings of polio reemergence and outbreaks of cholera, norovirus, Hepatitis A and Hepatitis B (Brouwer et al., 2018; Blomqvist et al. 2012; Hellmer et al., 2014; Madico et al., 1996). During the SARS epidemic in 2002, traces of SARS coronavirus were detected in wastewater near hospitals in China (Wang et al., 2005).

The first successful detection of SARS-CoV-2 in wastewater was reported in the Netherlands in early March 2020 (Medema et al., 2020); followed by demonstration of SARS-CoV-2 detection in wastewater in the United States (Wu et al., 2020; Sherchan et al., 2020). Wastewater surveillance for COVID-19 has since been broadly pursued by the scientific community, municipal public health and public works departments, and the US Centers for Disease Control and Prevention and other national organizing bodies in the United States and globally (Medema et al., 2020; Ahmed et al. 2020; Randazzo et al., 2020; Graham et al., 2020; La Rosa et al., 2020; Bivins et al., 2020; CDC NWSS).

Multiple applications of WBE for COVID-19 have been proposed, including as a leading indicator of new COVID-19 cases, an independent confirmation of disease trends, and as an early warning system for COVID-19 re-emergence (Olesen et al., 2021). Additionally, wastewater has been proposed as an alternative method to estimate the COVID-19 reproductive number (Huisman et al., 2021) or as an indicator of clinical diagnostic testing capacity (Xiao, Wu, Bushman et al., 2021). In practice, WBE has been applied in multiple ways. In Cambridge, MA, SARS-CoV-2 concentrations were one of three metrics used to evaluate school system re-openings (Salim 2020). In Australia, where COVID-19 cases are low, wastewater monitoring serves to warn residents of potential re-emergence (Gold Coast 2021). Across the United States, wastewater analysis has been deployed at universities to monitor and mitigate transmission among students (Harris-Lovett et al., 2021).

This study presents results from a nationwide dataset of SARS-CoV-2 concentrations in wastewater, and represents the largest US-based temporal and geographic WBE dataset reported to-date. In March 2020, our groups launched a campaign to monitor COVID-19 in wastewater across the United States (Wu et al., 2020; Biobot Analytics 2020; Wu, Xiao, Zhang et al., 2021). Since then, we have had approximately 100 participating locations regularly monitoring SARS-CoV-2 wastewater concentrations in their communities on a weekly or monthly basis. As a result of this effort, we have generated a wastewater SARS-CoV-2 dataset consisting of over 10,000 samples, allowing us to evaluate the applicability and feasibility of implementing a nationwide wastewater monitoring for COVID-19 disease. In this study, we report results from a subset of this data, highlighting data from 55 sampling locations with approximately weekly sampling for at least six months. These sampling locations represent 39 counties across 19 states, and we show their data from April 2020 through May 2021. We demonstrate that wastewater data broadly reflects geographic and temporal trends in COVID-19 cases across the United States, confirming the feasibility and applicability of wastewater monitoring for current and future public health COVID-19 disease surveillance.

## Methods

### Sample collection

The samples in this secondary research study were collected by participating wastewater treatment facilities as part of regular wastewater surveillance service provided by our company (Biobot Analytics, Inc; Cambridge, MA). Participating facilities were mailed a sampling kit with instructions to collect a standard 24-hour composite sample. Composite samples were shaken to homogenize and then aliquoted into three 50-mL conical tubes and shipped within 24 hours of collection overnight with ice packs to our laboratory (Cambridge, MA). Received samples were immediately pasteurized at 60°C for 1h. One of the three sample tubes was used for analysis, and processed immediately after pasteurization.

Samples were collected by the participating municipalities, who sampled at wastewater treatment facilities or pump stations. The majority of samples were collected using auto-samplers that these facilities already had in-house, including both refrigerated and non-refrigerated models. The majority of samples were collected as 24-hour composite samples. We encouraged our sampling partners to increase their auto-sampler’s pumping frequency to ensure representativeness of samples. Sampling frequency was determined by the participating municipal partners, with the majority providing weekly samples. Participating municipalities provided metadata about their sampling locations, including catchment population and average daily flow rate.

### Lab analysis

Our lab method changed over the sampling time course to improve sample processing time, throughput, and sensitivity, also accounting for supply-chain availability. In both methods, samples for analysis were first filtered to remove large particulate matter using a 0.2uM vacuum-driven filter (EMD-Millipore SCGP00525 or Corning 430320, depending on sample turbidity). Our initial lab method (“PEG-concentrated”) used PEG-salt precipitation to concentrate viruses from 40mL of wastewater, as described in Wu et al. (Wu et al., 2020). Resulting pellets were resuspended in TRIzol (ThermoFisher 15596026), and RNA was purified via phenol-chloroform extraction. RNA samples were first reverse transcribed (NEB M0368) and then assayed by qPCR (ThermoFisher 4444557) using SARS-CoV-2 nucleocapsid (N) gene (N1 and N2 regions), and PMMoV amplicons (CDC 2020; Zhang et al., 2006) on CFX96 or CFX-Connect instruments (BioRad).

In June 2020, we switched to our second method (“Amicon-concentrated”) which uses Amicon Ultra-15 centrifugal ultrafiltration units (Millipore UFC903096) to concentrate 15mL of wastewater approximately 100x. Viral particles in this concentrate are immediately lysed by adding AVL Buffer containing carrier RNA (Qiagen 19073) to the Amicon unit before transfer and >10 minute incubation in a 96-well 2mL block. To adjust binding conditions, 100% ethanol was added to the lysate, and samples were applied to RNeasy Mini columns or RNeasy 96 cassettes (Qiagen 74106 or 74181). For a subset of samples, we processed 30 mL of wastewater across two separate Amicon units, extracted each separately, and pooled the duplicate RNA extracts together prior to analysis. RNA samples eluted from the RNeasy kit were subjected to one-step RT-qPCR (ThermoFisher 4444436) analysis in triplicate for N1, N2, and PMMoV amplicons on CFX96 and/or CFX-Connect instruments. Cts were called from raw fluorescence data using the Cy0 algorithm from the qpcR package (v1.4-1) in R (Guescini et al., 2008), and manually inspected for agreement with the raw traces in the native BioRad Maestro software. Overall, we processed 667 samples with the first PEG concentration method; 3,497 samples with the second Amicon concentration method; and 11 samples with both (see discussion of sample reruns below).

In both methods, we ran a variety of laboratory controls. Positive synthetic SARS-CoV-2 RNA controls were included on every qPCR plate. Ct values for N1/N2 primers outside 31-33 triggered a plate re-run. Two no-template controls were included on every qPCR plate; N1/N2 Ct values > 40 and PMMoV Ct values > 35 for these controls triggered a plate re-run. One set of extraction blank controls was also run each day. Matrix inhibition was assessed manually by reviewing the raw qPCR curves. Finally, we used PMMoV as a proxy measure for per-plate recovery, and flagged any qPCR plates with unusually low PMMoV values for further review and potential plate re-run. Only results which passed all quality controls are reported here.

In addition to these laboratory controls, we implemented a thorough data review process, in which results were manually reviewed if they met certain additional criteria. These criteria included: PMMoV being below 1st or above 99th percentile of previously observed values; SARS-CoV-2 concentrations changing more than 5-fold since the prior sample; inhibition suspected from manual inspection of qPCR curves; concentrations obtained from N1 and N2 primers not concordant; and pigmentation present in extracted RNA. During the manual review process, we inspected individual replicates of qPCR and timelines of SARS-CoV-2 and PMMoV concentrations. A small fraction of samples that underwent the review process were flagged for a lab rerun. Reruns were done in duplicate if capacity allowed. If sufficient filtered wastewater from the initially-processed tube remained, a second aliquot from that tube was tested. We also always processed an aliquot using a second of the three 50 mL tubes of sewage. Approximately 2% of all samples were re-run through that process. If a rerun differed substantially from the original result, the original result was discarded (approximately 8% of reruns) and the rerun results were reported to the participating facility and included in this analysis; if the rerun recapitulated the original result, the average of all results were reported to the participating facility and included in this analysis.

### Data processing

A standard curve was generated using serial dilutions of Twist Bioscience synthetic SARS-CoV-2 RNA control 2 (MN908947.3) and used to convert Ct values into copies per well. We used pepper mild mottle virus (PMMoV) as a fecal indicator. Since synthetic RNA for PMMoV was not available, we used a DNA gBlock to build a standard curve for PMMV quantification.

Copies per well measured by qPCR were multiplied by a dilution factor accounting for the volume changes described above (RNA extraction, concentration, etc.) and then divided by the original sewage volume (40 mL, 30mL, or 15 mL) to convert to a sewage concentration. Non-detected wells were replaced with zero for these calculations. Concentrations of N1 and N2 replicates were averaged first within each primer set and then across primers to get the final SARS-CoV-2 concentration; replicates of the PMMoV amplicon were averaged. Samples were required to have at least two detected replicates between N1 or N2, and at least one detected PMMoV replicate to be considered a detection and subsequently quantified. To derive a normalized concentration, SARS-CoV-2 concentrations were divided by the PMMoV levels and multiplied by a reference PMMoV value derived as the median of our dataset comprising samples up to July 31, 2020 (3.65e8 copies/L). Results were returned to participating municipalities in the form of a PDF report containing the raw viral copies per liter of sewage and normalized concentrations, typically within 1 business day.

### Data analysis

We excluded locations representing fewer than 5000 people in their catchment population to reduce data variability, and locations which did not consent to data sharing. To evaluate the correlations between wastewater results and COVID-19 cases, we analyzed time series for all sampling locations with at least one sample per month for at least six months in the period of June 2020 through May 2021. Sampling location characteristics, including the population and associated county serviced by the catchment, were provided by treatment plant operators from each location (Table S1).

Daily COVID-19 cumulative case data was downloaded from USAFacts.org, which collated daily case counts for each US county from publicly available reports (USAFacts). To prevent negative new cases, any days with non-monotonically increasing cumulative case numbers (or with missing or non-numeric values) were replaced with the prior valid cumulative case number. New cases were then calculated as the difference in cumulative cases between consecutive days. Seven-day centered moving averages were calculated using the pandas .rolling() function (McKinney 2010). To derive incidence rates per 100k people, 7-day averages of reported cases were divided by the reported county populations, which were also downloaded from USAFacts.org (USAFacts).

Monthly averages for the maps were calculated as follows: for the SARS-CoV-2 virus concentration, we calculated the average normalized concentration per location for each month, and then averaged these values across locations within individual states. For the reported cases, we first calculated the per capita new cases by dividing daily reported cases by the county population and multiplying by 100,000. We then selected just the subset of counties represented in our selected sampling locations and averaged daily reported cases per capita over the month within each county, and then averaged across counties within the same state per month.

Statistical analyses and visualizations were performed in Python 3.8.

## Results

### Sampling locations

We selected sampling locations which had a suitably long time series starting from June 2020, when we started offering Covid-19 wastewater surveillance as a commercial service. Prior to June 2020, sampling locations participated pro-bono as part of an academic collaboration (Wu and Xiao, 2021). Fifty-five sampling locations collected at least one monthly sample for at least six months June 2020 through May 2021. These 55 locations represent approximately 12,500,000 people (mean = 245,000 and median = 55,000 people per location; Table S1, Fig S1) distributed across 39 counties in 19 states, with a maximum of 5 locations in one county. These locations took an average of 4.4 samples per month (Table S1, Fig S2).

### Temporal trends

We compared the trends in wastewater virus concentration with reported cases in the respective counties for the 55 sampling locations. Wastewater SARS-CoV-2 concentrations closely followed the 7-day average of new clinical cases (Fig 1), correlating well in the majority of locations (median Spearman correlation = 0.75, IQR = 0.65 to 0.83; Fig S3). Wastewater measurements mirrored a rapid increase in clinical COVID-19 cases in late October through November 2020 that occurred in every county we sampled. Of note, many locations experienced the highest viral concentrations and highest clinical case counts observed to-date in their November 2020 through January 2021 samples, reflecting the COVID-19 winter resurgence experienced nationwide in the US (Figs 1 and 2). Wastewater also reflected the decrease in clinical cases in the first months of 2021 (Fig 1).

**Figure 1.**
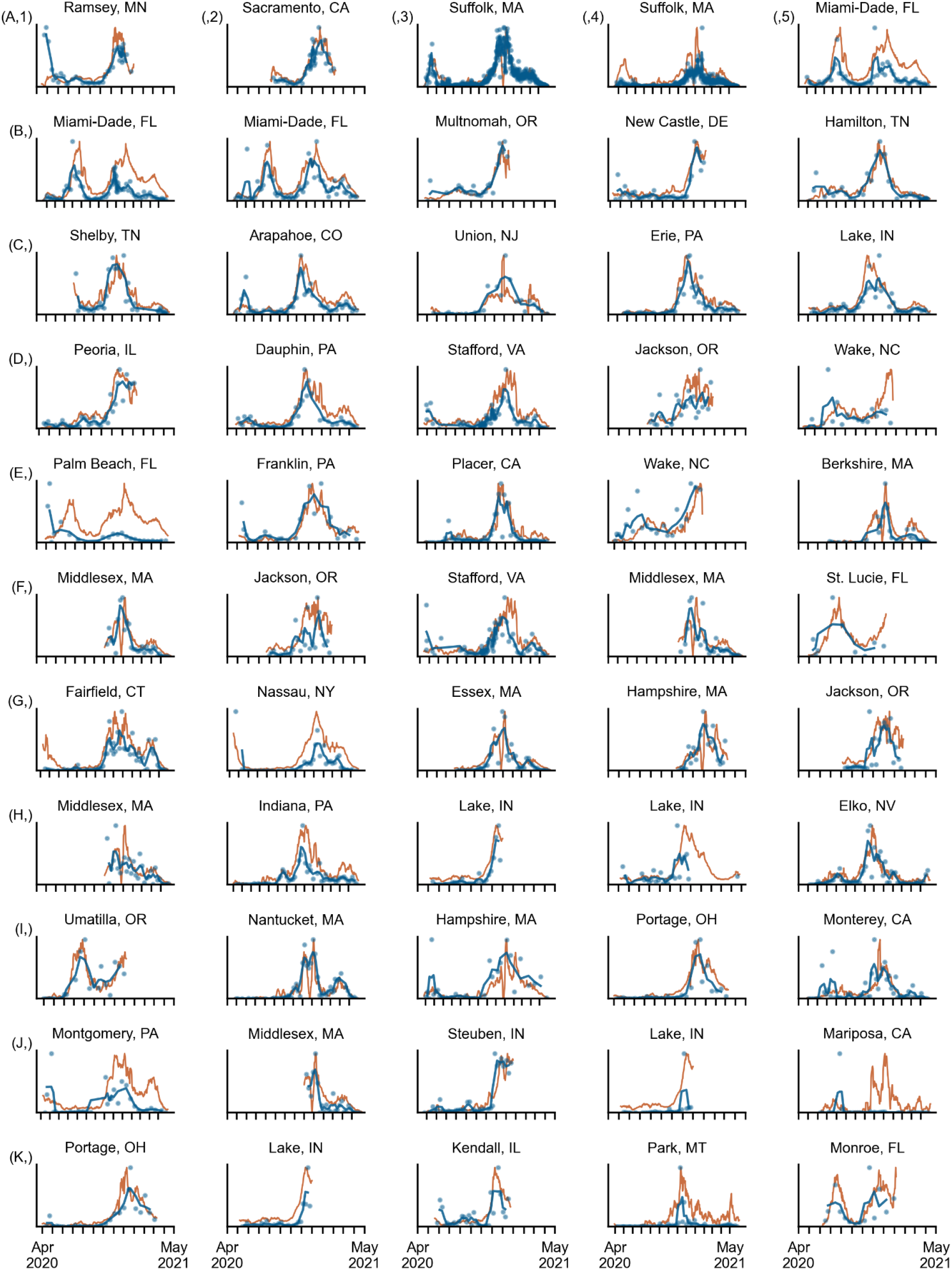
Time series of results from sampling locations with at least one monthly sample for at least six months June 2020 through May 2021 (n=55). Blue line: centered 3-sample average of normalized wastewater concentrations (genome copies/L); blue dots: individual normalized wastewater concentration measurements (genome copies/L); orange line: centered 7-day average of daily new cases in the respective county (new cases). Y-axes are normalized to the maximum of each time series and lines are shown without units to emphasize relative trends within each location. X-axes are consistent across plots, with monthly ticks ranging from April 1st, 2020 to May 1st, 2021. Plots are labeled with county and state names of the associated catchment. Sampling locations are organized in order of decreasing catchment population; i.e. the largest catchment (1,950,000 people) is at the top left (A1) and the smallest catchment (6,400 people) is the bottom-right-most plot (K5). Rows are labeled with letters and columns with numbers for ease of reference. Individual time series for each location, including daily new cases and detailed units for both axes, are provided in Supplementary File 2.

**Figure 2.**
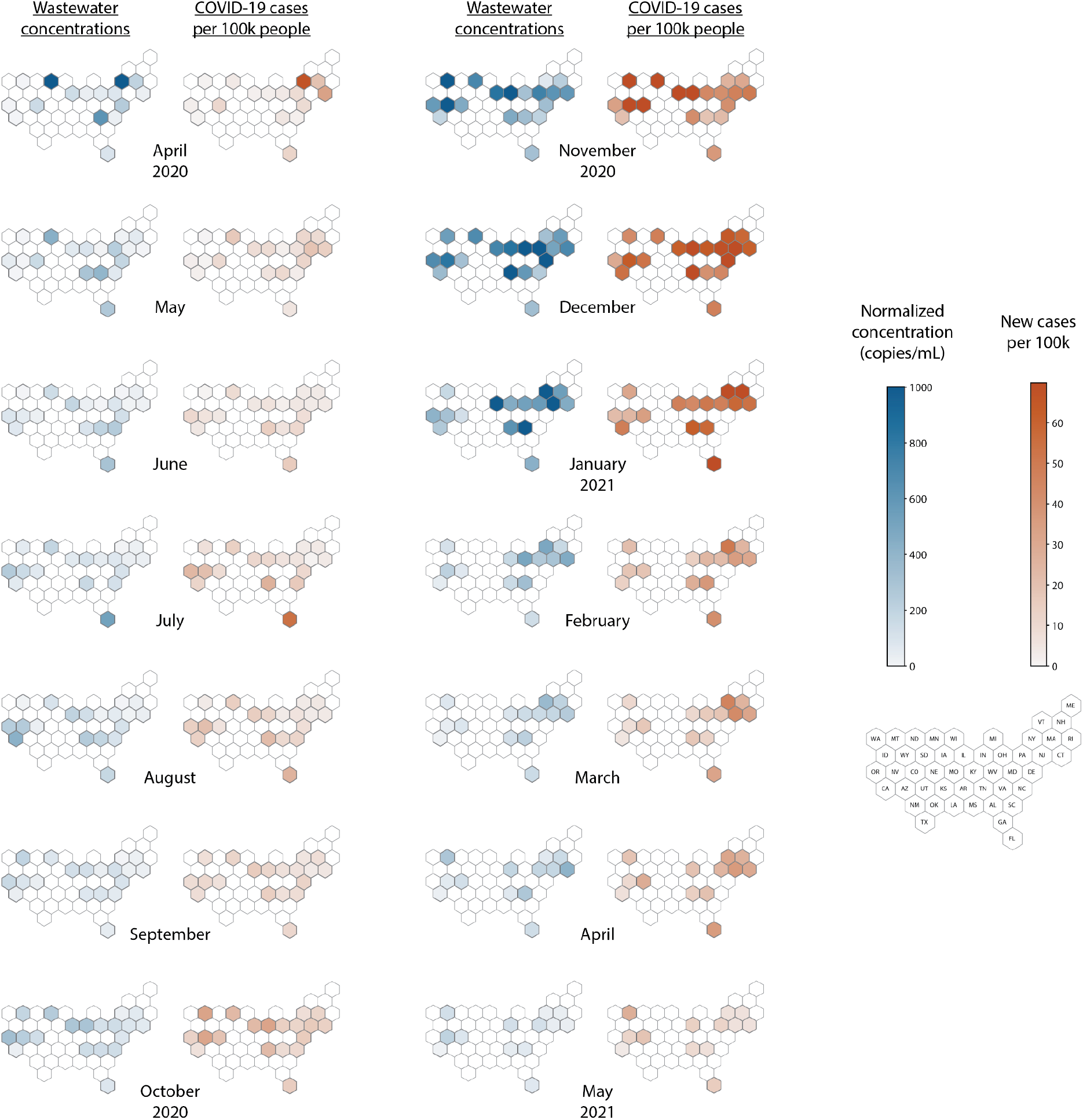
Monthly geographic trends in wastewater SARS-CoV-2 concentrations and COVID-19 cases. (Blue data; first and third columns from the left) Monthly state-level normalized wastewater SARS-CoV-2 concentrations. Monthly state average is calculated as the average normalized concentration per sampling location per month, then averaged across sampling locations within the same state. (Orange data; second and fourth columns from the left) Monthly state-level new COVID-19 cases per 100,000 people for counties with at least one wastewater sampling location in that month. Monthly state average is calculated as the monthly average of new daily cases per 100,000 people in counties with a corresponding sampling location, then averaged across counties within the same state. States which did not include any sampling locations are outlined in gray. Color scales apply to all maps and are truncated at the 95th percentile of each dataset.

Wastewater also tracked location-specific qualitative trends throughout the year. For example, a subset of counties experienced peaks in COVID-19 cases in the summer (June-August 2020) which were also well-tracked by wastewater data. For example, wastewater concentrations in all three sampling locations in Miami-Dade County, FL (Fig 1, A5, B1, B2) reliably reflected the two distinct waves of Covid-19 that Florida experienced during the sampling period. Cases and wastewater concentrations both increased in summer 2020, decreased to a new baseline, increased again in late 2020, and decreased once more in early 2021. Similar patterns were observed in the other smaller Florida sampling locations (Fig 1, E1, F5, and K5). Counties in other states experienced COVID-19 peaks earlier in 2020. For example, Suffolk County, MA experienced a peak in COVID-19 cases in April 2020, which was reflected in the wastewater data from both associated sampling locations (Fig 1, A3-4). A similar trend was observed in Hampshire County, MA (Fig. 1, I3). Finally, wastewater reflected different dynamics within the decreasing clinical cases early in 2021. For example, in Suffolk County, MA, wastewater seemed to plateau in February and March 2021 before decreasing again in April, a trend which was also seen in clinical cases. Similarly, many locations (Stafford County, VA (D3 and F3); Arapahoe County, CO (C2); Dauphin, PA (D2); Erie, PA (C4); Lake, IN (C5)) had a slight uptick in clinical cases around April 2021 which was reflected by the wastewater measurements. The ability of wastewater to track these more nuanced epidemiological dynamics supports its ability to provide independent confirmation of relative disease burden and trends (Fig 1, Supplementary File 2; Olesen et al., 2021).

We observed a range of correlations between wastewater measurements and new clinical cases across the 55 sampling locations (Fig S3, Table S2). We first explored whether the correlation strength was associated with external factors, and found no relationship between the strength of the correlation in a sampling location and its population, average reported flow, sampling frequency, or county coverage (Fig S4). Interestingly, the lack of relationship with county coverage indicates that precise geographical alignment may not be required to evaluate broad trends in wastewater measurements when sampling at the municipality level.

Next, we explored the impact of normalizing raw SARS-CoV-2 concentrations with a human fecal marker (PMMoV) on the observed correlations. We found that normalizing by PMMoV did not always improve the correlation between wastewater measurements and reported clinical cases within individual time series (Fig S3, Table S2). In some locations, normalization improved the correlation but in others raw virus concentrations correlated better with cases (Fig S3.B). These results confirm findings by others who have investigated the impact of normalization in multi-site studies and found no clear impact of normalization on correlations (Feng et al., 2021; Sweetapple et al. 2021). We hypothesize that the broad range of locations profiled lead to large variability in wastewater matrices and therefore similarly large variability in how much impact normalization has on the data quality.

### Geographic trends

We next compared geographic trends in average monthly SARS-CoV-2 wastewater concentrations and reported COVID-19 cases across states (Fig 2, Fig S5, Table S3). Overall, wastewater concentrations showed similar relative patterns across states as did clinical cases (median Spearman correlation across states = 0.6; IQR=0.52 to 0.81). These results indicate that normalized wastewater concentrations can be used to compare relative Covid-19 burden across different geographies.

The strength of the geographic correlation varied throughout the course of the pandemic. The second-lowest correlation was observed early in the pandemic in May 2020 (Spearman correlation = 0.4), which could be explained in part by different testing capacities across states, leading to geographic differences in the completeness of the case data. Similarly, a lower correlation in December 2020 could have been affected by irregularities in case reporting around the American winter holidays. In contrast, some of the highest correlations between wastewater and clinical cases across states were observed in the summer 2020 wave, when Covid-19 was rapidly increasing and clinical testing was well-established nationwide (June 2020= 0.79; July = 0.90; August = 0.86) (Fig S5, Table S3).

We next investigated the effect of normalization on the ability for wastewater concentrations to reflect patterns in clinical cases across locations. We found that raw wastewater concentrations across states did correlate with cases in most months, but that these correlations were weaker than when looking at the normalized concentration (Fig S6, Table S3). In fact, normalization improved the geographic correlation in all but two months (Fig S7.B).

### Lessons learned from nationwide implementation

Through this work, we identified several key practical considerations for implementing a long-term nationwide wastewater monitoring program. First, the impact of changing laboratory methods on overall data interpretation was minimal. From a scientific standpoint, changing methods throughout the course of a study is usually not recommended due to the possibility of introducing batch effects. However, this secondary research study uses a dataset generated through a regular commercial wastewater surveillance service, which required that the laboratory methods changed as our experience improved and reagent supply chains stabilized (Methods). We investigated the impact of these method changes on our dataset, and found that the overall interpretation of the data remained unaffected (Figs S8-9). Importantly, updating methods to reflect current scientific understanding and changing epidemiological contexts will be required for any nationwide WBE monitoring system implemented in practice. For example, as COVID-19 cases decrease substantially, methods will need to be updated to improve sensitivity. The results presented here demonstrate agreement between wastewater measurements and clinical cases despite methodological changes, highlighting that while addressing the impact of variations in laboratory methods remains an important area of scientific inquiry, it is not necessary to have a gold standard methodology established for wastewater-based epidemiology to be implemented at scale and to provide reliable reflections of public health trends. From our experience, the most necessary component to address the impact of method changes was establishing collaborations and trust between academics, wastewater and public health officials, and groups deploying WBE at scale.

Second, we found that identifying best practices for wastewater-based epidemiology requires a holistic approach beyond purely scientific considerations. For example, rapid turnaround time of the data quickly emerged as a key requirement for our data to be useful to our sampling partners and their public health counterparts. Therefore, our method development efforts prioritized minimizing the operational impact to turnaround time above other considerations. With these considerations, any changes to our methods and data interpretation required careful considerations of data continuity and communication to relevant stakeholders. Moreover, we found that a manual data review process was essential for ensuring quality of the data, development of QC metrics, and understanding of the data trends. As WBE expands in practice, difficult trade-offs will need to be made when improving data quality conflicts with operational requirements.

Finally, logistical considerations also dictate which datasets can be used as epidemiological comparisons for WBE at scale. For example, precise geographic alignment between catchment sewersheds and reported cases was not feasible, as it would have required requesting and coordinating GIS data across a different set of municipal partners for each sampling catchment. We used data provided by USA Facts for our clinical comparison because at the time we launched this work, it was the only data source that provided geographical comparisons more granular than state level and which was licensed appropriately for our use. Future WBE efforts implemented at scale will similarly need to leverage systematically collected and curated third-party datasets or expend significant resources to compile them themselves. Importantly, clinical data itself is not necessarily a gold standard for comparing wastewater-based results. In the case of COVID-19, reported cases are used as a benchmark to evaluate how well wastewater is reflecting broad trends, but are not necessarily comprehensive due to asymptomatic infections and limitations or inequalities in access to diagnostic testing (Olesen et al., 2021). In fact, the comprehensiveness of wastewater is one of its key strengths, and deviations between wastewater and clinical data may reflect limitations in the clinical data itself (Xiao, Wu, Bushman, et al., 2021).

## Discussion

Effective and long-term systems will be required to continuously monitor SARS-CoV-2 across the United States. While diagnostic testing and clinical case reporting continues to drive public health recommendations, wastewater assessment of SARS-CoV-2 is a practical and sustainable complement that can be integrated into existing wastewater infrastructure to provide cost-effective population-level monitoring. With effective vaccines against SARS-CoV-2, persistent surveillance to detect resurgences of the virus will become increasingly important. Here, we demonstrate that wastewater data reflects both temporal and geographic trends in COVID-19 disease burden across the United States, suggesting its utility on a national scale as a SARS-CoV-2 surveillance system.

Our study has several limitations. First, wastewater SARS-CoV-2 concentrations are highly variable due to sampling, lab processing, and qPCR analysis. We attempted to reduce this challenge through normalization with a fecal virus with biological similarities to SARS-CoV-2 (Wu et al., 2020) and by optimizing our lab protocols (Methods). However, we still observed large spikes in wastewater concentrations in some locations which were not explained by changes in reported COVID-19 cases. These data challenges are common in the field of wastewater-based epidemiology, and our group and others are actively developing models to correct for variability and better interpret spikes. Second, the geographic comparisons between wastewater results and reported cases are not exact, as sampling location catchments may represent a subset or superset of the respective comparison counties (Table S1). However, we found no relationship between a catchment’s coverage of its comparison county and its correlation between wastewater and cases, indicating that these wastewater catchments were sufficient to identify broad and general trends in their associated communities (Fig S4). Alternative public health applications of wastewater-based epidemiology like building-level or manhole sampling may require more granular insight, in which geographies may need to be more precisely aligned. Third, wastewater data showed a range of correlation strengths with reported cases (Fig 1, Fig S3). In addition to technical and geographic factors, these differences could also be due to different testing capacity across space and time. For example, access to testing was extremely limited early in the pandemic, which may in part explain the lower geographic correlation at that time.

Despite these limitations, wastewater assessment of SARS-CoV-2 reliably reflected trends in clinical cases within and across locations, confirming the feasibility of acquiring useful data from a broad variety of wastewater facilities. The sampling locations profiled in this study were selected in a non-biased fashion without any pre-consideration of the wastewater results; we imposed no exclusion criteria beyond a catchment size cutoff of 5,000 people and a suitably long sampling period for the analysis. Furthermore, wastewater data reflected clinical trends across a range of wastewater facility characteristics, including catchment populations ranging from just above 6,000 people to close to 2 million individuals. Results in this study are comparable to prior work which has profiled individual sampling locations and found a range of correlation strengths between wastewater SARS-CoV-2 concentrations and clinical cases (Peccia et al., 2020; Stadler et al., 2020; Wu et al., 2020; Ahmed et al., 2020b). Deploying WBE to this large number of communities demonstrates the generalizability of these results on a large nationwide scale (CDC NWSS).

This study also provides additional insight into whether and how normalization impacts wastewater data interpretation. When analyzing wastewater concentrations as a reflection of Covid-19 trends within individual sampling locations, we found that normalization improved the correlation between wastewater and reported cases in some sampling locations, but not in others (Fig S3). In contrast, we found that normalization did improve correlations when comparing *across* locations (Fig S7). Finally, in our experience, fecal normalization had additional practical benefits beyond improving correlations, addressing concerns related to sample quality and potential dilution and serving as an additional endogenous laboratory control for all samples.

This work also demonstrates the practical feasibility of implementing and scaling a national wastewater surveillance system for COVID-19. Wastewater surveillance is straightforward to implement with the participation of wastewater utilities; we achieved broad uptake among different municipalities across the United States (Biobot Analytics 2020, Wu and Xiao, 2021). All of our participating municipalities were able to reliably collect samples using standard wastewater sampling devices and often as part of their regular sampling schedules, requiring little extra work on their part. Additionally, samples were sent to us through traditional mail services and the majority passed our internal quality control process (Methods). This confirms that national wastewater surveillance programs like those supported by the United States Centers for Disease Control and Prevention are logistically and practically feasible (CDC NWSS). With sufficient resources, a national WBE dashboard could complement similar population level mapping of SARS-CoV-2 based on clinical reports (Dong et al., 2020, Naughton et al., 2021), lending insights on key operational decisions like phased reopenings, geographic selection of testing locations, and hospital preparedness.

Overall, this work demonstrates that wastewater can be leveraged as a passive population-level surveillance strategy for COVID-19, and that implementing a nationwide monitoring system is feasible and sustainable. The dataset reported here is the largest US-based wastewater dataset for COVID-19 reported to date, and demonstrates that wastewater concentrations of SARS-CoV-2 reflect trends in COVID-19 burden in associated communities across a broad variety of sampling locations. Our ability to rapidly develop, deploy, and improve methods to detect SARS-CoV-2 in wastewater and our success in working with a large number of wastewater utilities further provides scientific and practical insights which can be used to support implementing wastewater-based epidemiology across the US as a standard component of the public health toolkit for COVID-19 and future infectious diseases.

## Data Availability

An anonymized version of the data is available at https://github.com/biobotanalytics/covid19-wastewater-data

## Acknowledgements

This work could not have been possible without the support of the participating wastewater utilities; we thank the sampling operators and treatment plant facilities for their efforts and collaboration as we established and scaled our processes, and for agreeing to this secondary research use of their data. We thank Jennings Heussner, Nicholas Santos-Powell, and the rest of the Biobot Analytics, Inc. and MIT Alm lab teams for logistical support. We also thank Genevieve Hoffman for support on the figure visual designs.

## Notes

**Conflict of interest and funding sources** C.D., K.A.M., N.E., M.I., R.F.O., M.M.P., S.M., S.T.W., F.C., T.M., C.L.S., and S.W.O are current or former employees of Biobot Analytics Inc. E.J.A. is scientific advisor to Biobot Analytics, Inc. N.G. and M.M. are co-founders of Biobot Analytics. P.R.C. is funded by NIH K23DA044874; P.R.C. and M.M. are funded by NIH R44DA051106; P.R.C., M.M., T.B.E. and E.J.A. are funded by the Massachusetts Consortium on Pathogen Readiness (MassCPR) and China Evergrande Group.

### Competing Interest Statement

C.D., K.A.M., N.E., M.I., R.F.O., M.M.P., S.M., S.T.W., F.C., T.M., C.L.S., and S.W.O are current or former employees of Biobot Analytics Inc. E.J.A. is scientific advisor to Biobot Analytics, Inc. N.G. and M.M. are co-founders of Biobot Analytics. P.R.C. is funded by NIH K23DA044874; P.R.C. and M.M. are funded by NIH R44DA051106; P.R.C., M.M., T.B.E. and E.J.A. are funded by the Massachusetts Consortium on Pathogen Readiness (MassCPR) and China Evergrande Group.

### Funding Statement

C.D., K.A.M., N.E., M.I., R.F.O., M.M.P., S.M., S.T.W., F.C., T.M., C.L.S., and S.W.O are current or former employees of Biobot Analytics Inc and received compensation as part of this work. E.J.A. is scientific advisor to Biobot Analytics, Inc. N.G. and M.M. are co-founders of Biobot Analytics. P.R.C. is funded by NIH K23DA044874; P.R.C. and M.M. are funded by NIH R44DA051106; P.R.C., M.M., T.B.E. and E.J.A. are funded by the Massachusetts Consortium on Pathogen Readiness (MassCPR) and China Evergrande Group.

