## Supplemental File 1 for "Nationwide trends in COVID-19 cases and SARS-CoV-2 wastewater concentrations in the United States"

* These authors contributed equally.

### Supplementary File 1: Supplementary Figures and Tables

#### Supplementary Figures

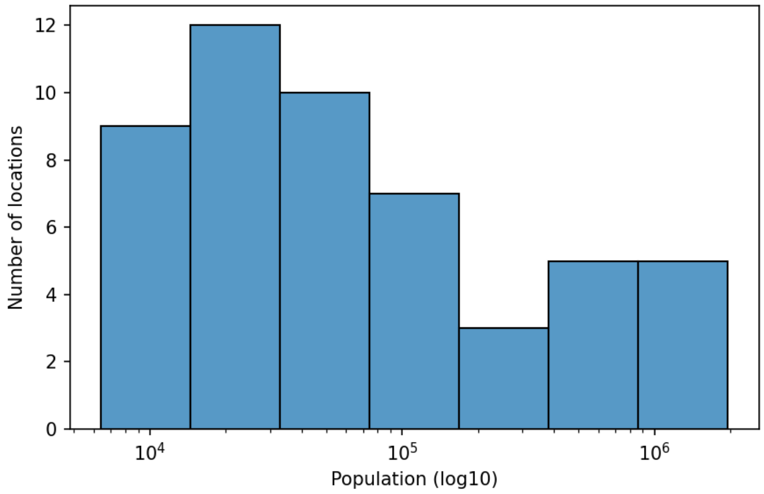


Figure S1. Distribution of catchment populations for all 55 locations profiled. Catchment populations were provided directly by the wastewater utilities and may represent actual population served or design capacity.

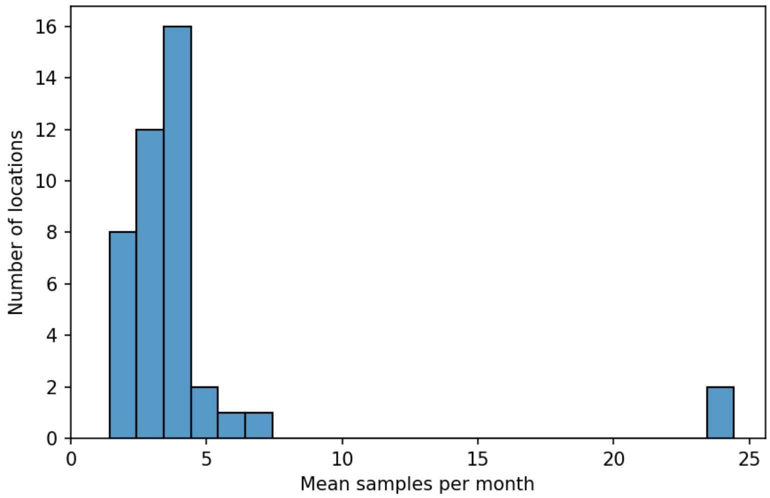


Figure S2. Distribution of sampling frequencies per location. Average sampling frequency was calculated by taking the average of the number of samples per month for each location, for all months that the location had at least one sample in.

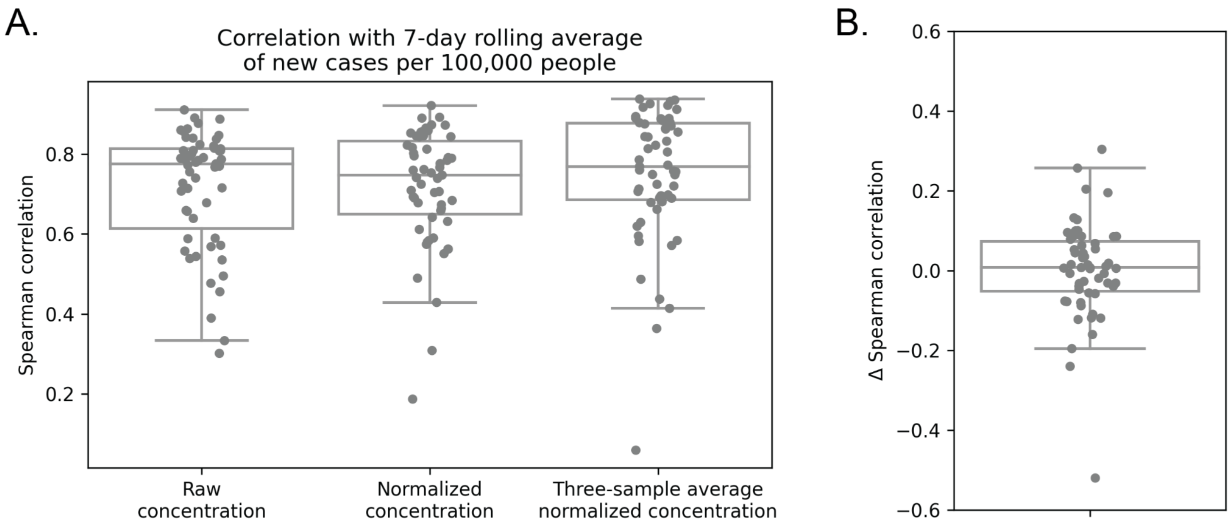


Figure S3. Temporal correlations between wastewater concentrations and new reported COVID-19 cases within locations. (A) Boxplot shows the Spearman correlation between wastewater SARS-CoV-2 concentrations (copies/L) and new reported clinical cases (total cases). Each point is the correlation for one location. Correlations were calculated using three different measures of wastewater concentration: raw, unadjusted SARS-CoV-2 concentration, SARS-CoV-2 concentrations normalized to a fecal maker (PMMoV), and a three-sample average of the normalized concentrations. (B) Difference in the Spearman correlation when calculated using normalized or raw concentrations. Each point is a sampling location; a positive delta indicates that the Spearman correlation calculated using normalized concentrations is higher than the correlation calculated using raw concentrations.

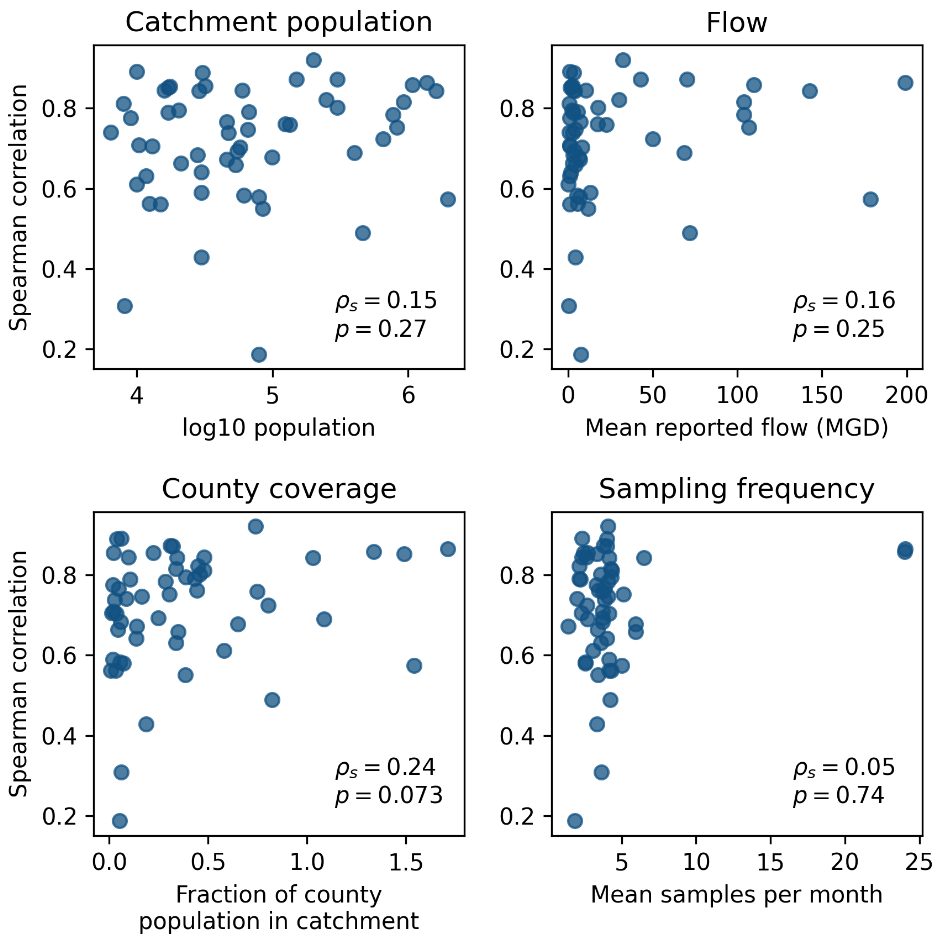


Figure S4. Relationship between temporal correlation and catchment characteristics. Each subplot shows the relationship between the Spearman correlation of wastewater concentrations and COVID-19 cases per location (y-axis) and one catchment characteristic (x-axis). Spearman correlations were calculated between each location's temporal correlation and its associated factor, using Python's `scipy.spearmanr` function. Spearman correlations and associated uncorrected p-values are reported on each plot. (Top row) Catchment population and flow were reported by the wastewater utilities. If a flow was provided for the day of sampling, that value was used, otherwise the reported yearly average flow was used for that sample. Flows were averaged across all samples per location. (Bottom row, left) County coverage was calculated as the catchment population (provided by the wastewater utilities) divided by the population of the primary county served by the wastewater facility. (Bottom row, right) Average monthly sampling frequency calculated per location. Correlation calculated without the two highest values is $r_s$ = -0.05; p = 0.73.

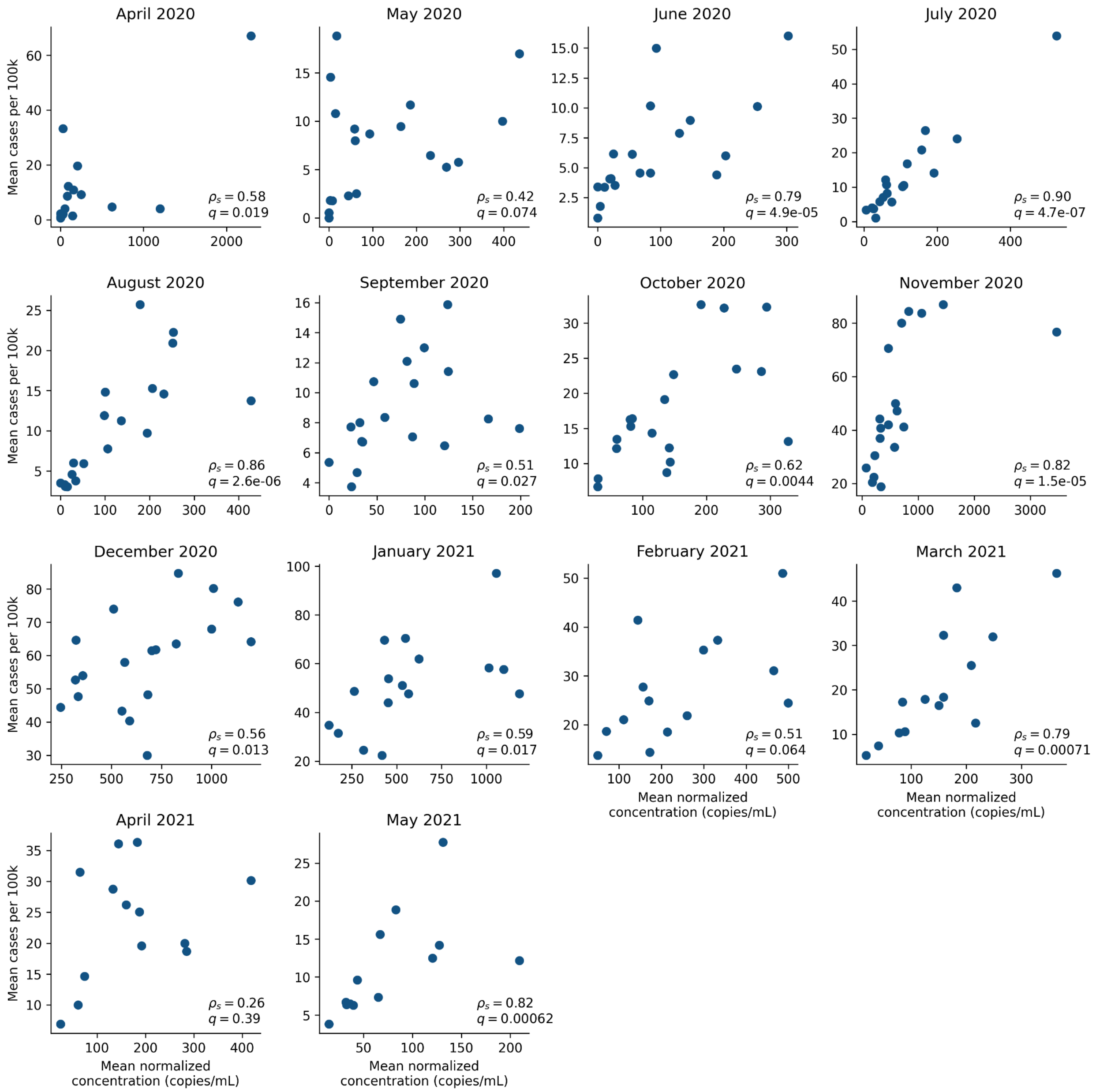


Figure S5. Geographic correlations per month. Scatterplots showing normalized concentrations (x-axis) vs. new cases per 100k (y-axis), averaged within each state (points) per month (subplots). Spearman correlations were calculated using `scipy.spearmanr`; correlations and uncorrected p-values are provided on each plot.

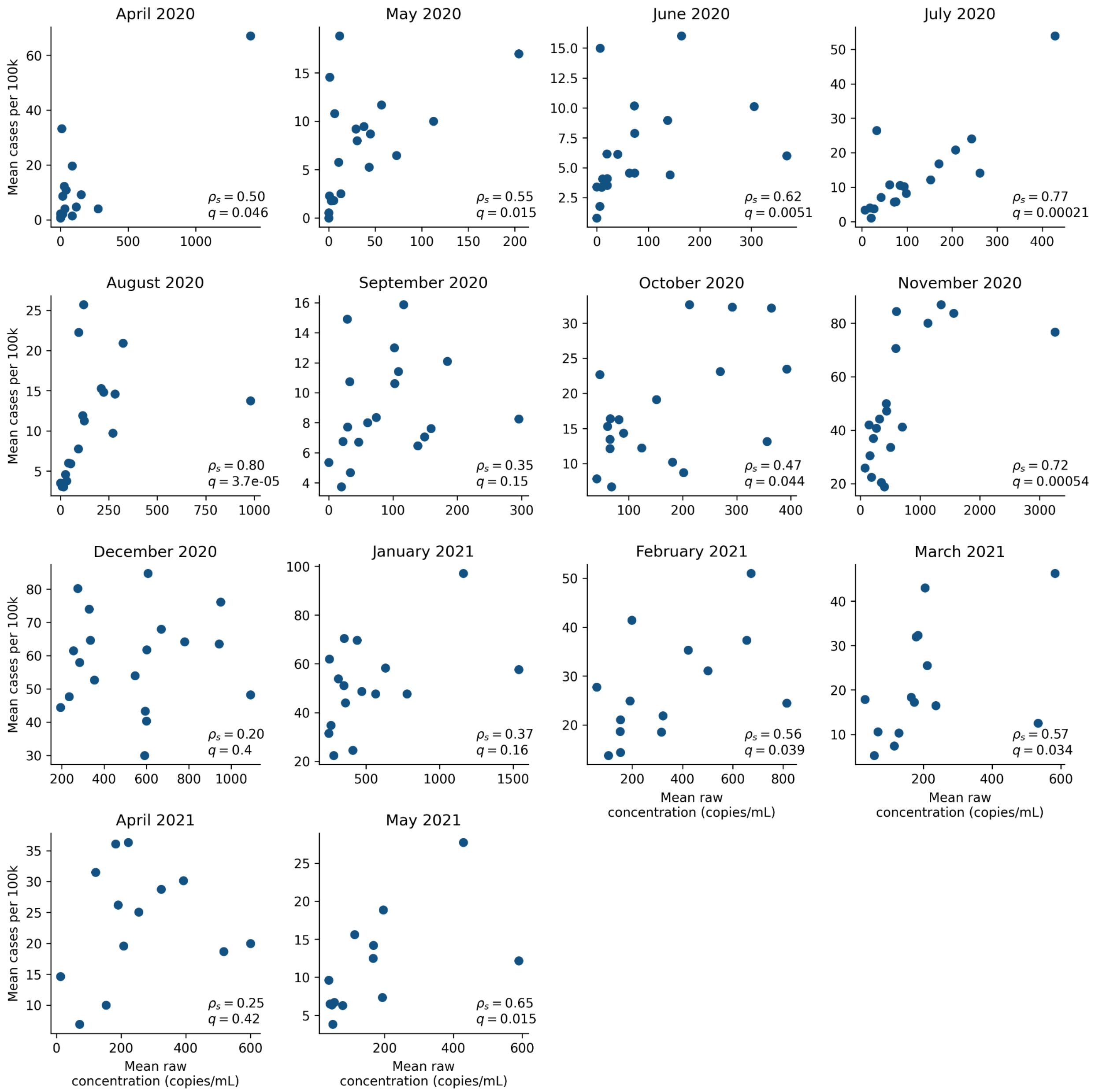


Figure S6. Same geographic correlations as above but using the monthly average of the raw concentrations instead of normalized concentrations.

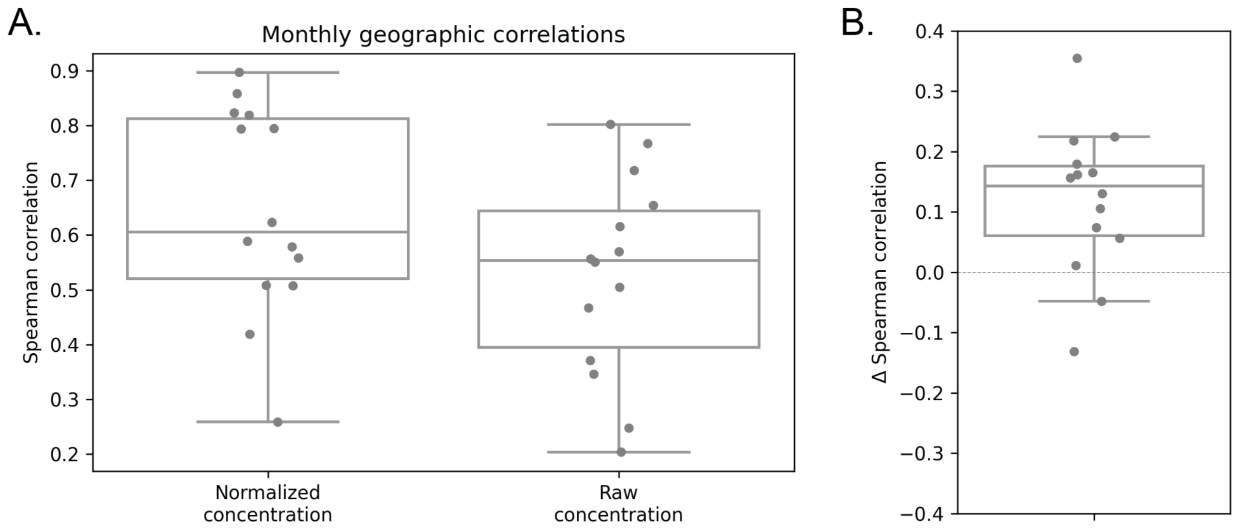


Figure S7. Geographic correlations with normalized vs. raw concentrations. (A) Each point is a month; y-axis is the correlation between states during that month. X-axis indicates which concentration measure was used to calculate the correlation. (B) Difference in the Spearman correlation when calculated using normalized or raw concentrations. Each point is a month; a positive delta indicates that the Spearman correlation across states calculated using normalized concentrations is higher than the correlation calculated using raw concentrations.

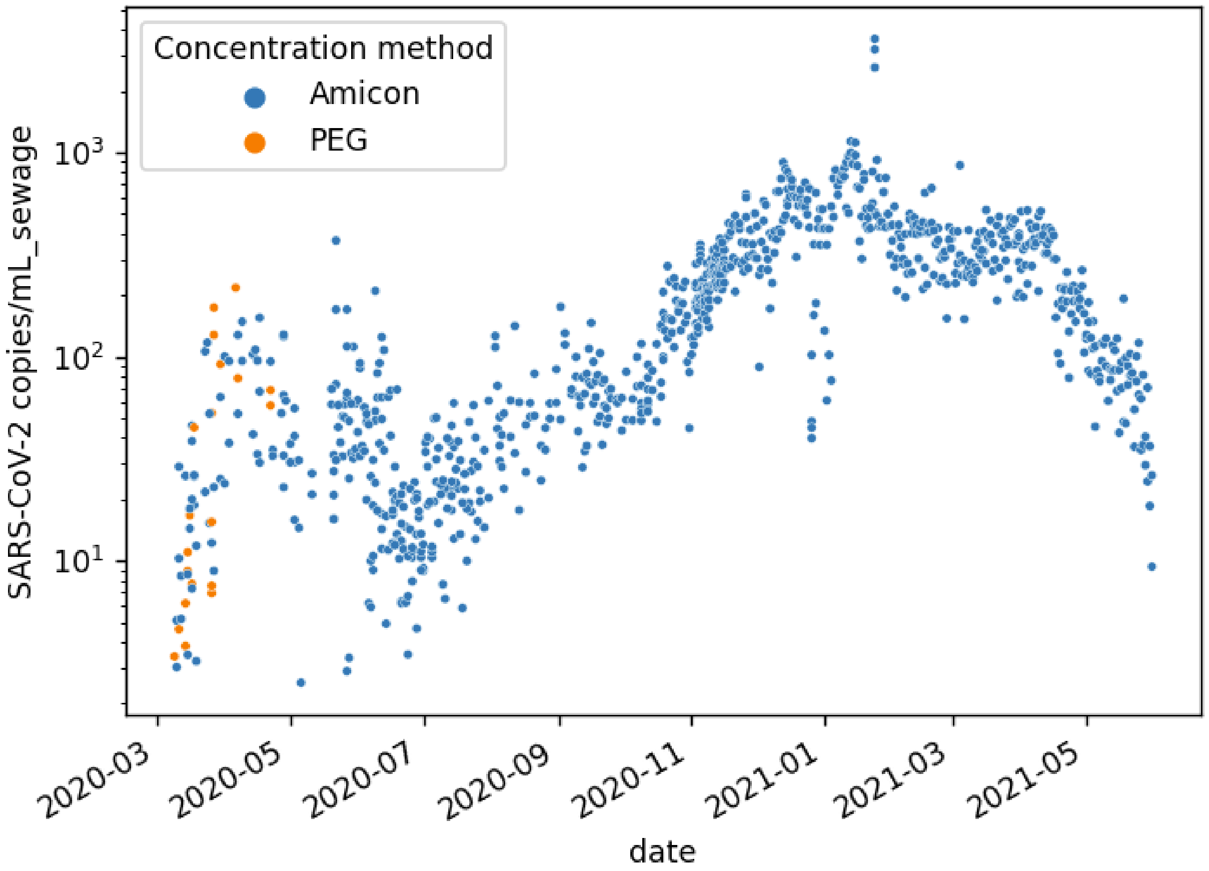


Figure S8. Time series of SARS-CoV-2 concentration in sewage (copies/mL) for the two Suffolk County, MA locations, with PEG-concentrated samples shown in orange, and Amicon-concentrated samples shown in blue (Methods).

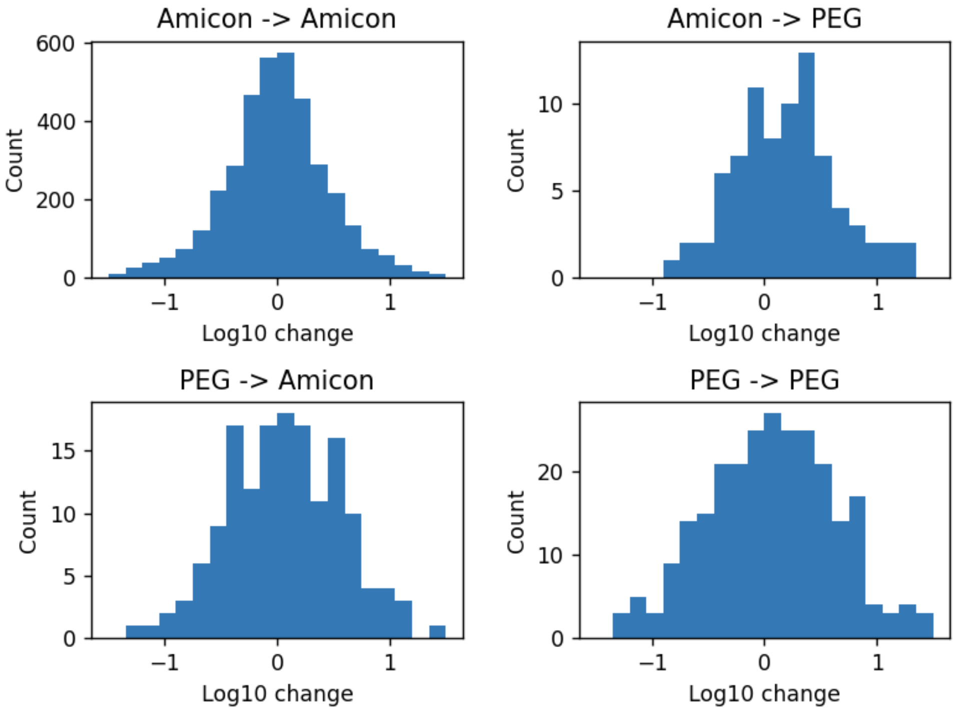


Figure S9. A histogram of changes between Amicon-concentrated and PEG-concentrated samples (Methods). A change was defined as a pair of samples in which a second sample was taken 0-7 days after the first one. For example, if a sample from 6/10 was concentrated with PEG, and another sample that was taken on 6/10 through 6/17 including 6/17 was concentrated with Amicon, this pair of samples would contribute to the histogram labelled “PEG -> Amicon”.

### Supplementary Tables

| Figure 1 label | County | State | Catchment population | Fraction of county covered | County population | Mean reported flow | Mean monthly sampling frequency |
|---|---|---|---|---|---|---|---|
| A1 | Ramsey | MN | 1,950,000 | 1.54 | 1,265,843 | 178.73 | 5.00 |
| A2 | Sacramento | CA | 1,600,000 | 1.03 | 1,552,058 | 142.82 | 6.50 |
| A3 | Suffolk | MA | 1,375,228 | 1.71 | 803,907 | 199.33 | 24.07 |
| A4 | Suffolk | MA | 1,075,656 | 1.34 | 803,907 | 109.75 | 24.00 |
| A5 | Miami-Dade | FL | 920,528 | 0.34 | 2,716,940 | 103.97 | 4.29 |
| B1 | Miami-Dade | FL | 829,725 | 0.31 | 2,716,940 | 106.70 | 5.13 |
| B2 | Miami-Dade | FL | 776,150 | 0.29 | 2,716,940 | 104.01 | 4.07 |
| B3 | Multnomah | OR | 654,741 | 0.81 | 812,855 | 50.13 | 2.67 |
| B4 | New Castle | DE | 460,000 | 0.82 | 558,753 | 71.80 | 4.22 |
| B5 | Hamilton | TN | 400,000 | 1.09 | 367,804 | 68.73 | 2.69 |
| C1 | Shelby | TN | 300,000 | 0.32 | 937,166 | 70.36 | 4.00 |
| C2 | Arapahoe | CO | 300,000 | 0.46 | 656,590 | 17.81 | 3.60 |
| C3 | Union | NJ | 250,000 | 0.45 | 556,341 | 30.00 | 2.15 |
| C4 | Erie | PA | 200,000 | 0.74 | 269,728 | 32.33 | 4.08 |
| C5 | Lake | IN | 149,941 | 0.31 | 485,493 | 42.97 | 3.77 |
| D1 | Peoria | IL | 134,000 | 0.75 | 179,179 | 22.70 | 3.70 |
| D2 | Dauphin | PA | 124,000 | 0.45 | 278,299 | 17.30 | 3.46 |
| D3 | Stafford | VA | 99,373 | 0.65 | 152,882 | 6.25 | 5.92 |
| D4 | Jackson | OR | 85,000 | 0.38 | 220,944 | 12.01 | 3.43 |
| D5 | Wake | NC | 79,500 | 0.07 | 1,111,761 | 7.06 | 2.56 |
| E1 | Palm Beach | FL | 79,100 | 0.05 | 1,496,770 | 7.48 | 1.85 |
| E2 | Franklin | PA | 67,000 | 0.43 | 155,027 | 5.69 | 2.17 |
| E3 | Placer | CA | 66,000 | 0.17 | 398,329 | 4.43 | 4.07 |
| E4 | Wake | NC | 61,700 | 0.06 | 1,111,761 | 5.38 | 2.56 |
| E5 | Berkshire | MA | 60,000 | 0.48 | 124,944 | 10.61 | 2.64 |
| F1 | Middlesex | MA | 57,909 | 0.04 | 1,611,699 | 8.16 | 4.14 |
| F2 | Jackson | OR | 55,000 | 0.25 | 220,944 | 3.84 | 3.71 |
| F3 | Stafford | VA | 53,508 | 0.35 | 152,882 | 4.30 | 5.92 |
| F4 | Middlesex | MA | 47,152 | 0.03 | 1,611,699 | 2.64 | 3.86 |
| F5 | St. Lucie | FL | 46,000 | 0.14 | 328,297 | 6.96 | 1.43 |

| | | | | | | | |
|---|---|---|---|---|---|---|---|
| G1 | Fairfield | CT | 45,878 | 0.05 | 943,332 | 7.37 | 3.86 |
| G2 | Nassau | NY | 32,000 | 0.02 | 1,356,924 | 2.77 | 2.71 |
| G3 | Essex | MA | 30,430 | 0.04 | 789,034 | 3.30 | 4.00 |
| G4 | Hampshire | MA | 30,000 | 0.19 | 160,830 | 4.41 | 3.33 |
| G5 | Jackson | OR | 30,000 | 0.14 | 220,944 | 1.90 | 4.00 |
| H1 | Middlesex | MA | 29,927 | 0.02 | 1,611,699 | 13.07 | 4.14 |
| H2 | Indiana | PA | 28,809 | 0.34 | 84,073 | 4.03 | 4.14 |
| H3 | Lake | IN | 28,040 | 0.06 | 485,493 | 3.04 | 3.71 |
| H4 | Lake | IN | 21,228 | 0.04 | 485,493 | 2.86 | 3.38 |
| H5 | Elko | NV | 20,467 | 0.39 | 52,778 | 2.56 | 4.31 |
| I1 | Umatilla | OR | 17,500 | 0.22 | 77,950 | 2.06 | 2.44 |
| I2 | Nantucket | MA | 17,000 | 1.49 | 11,399 | 1.43 | 3.36 |
| I3 | Hampshire | MA | 17,000 | 0.11 | 160,830 | 3.02 | 2.21 |
| I4 | Portage | OH | 16,028 | 0.10 | 162,466 | 3.06 | 2.33 |
| I5 | Monterey | CA | 15,000 | 0.03 | 434,061 | 1.16 | 4.31 |
| J1 | Montgomery | PA | 13,000 | 0.02 | 830,915 | 1.24 | 2.31 |
| J2 | Middlesex | MA | 12,397 | 0.01 | 1,611,699 | 5.70 | 4.17 |
| J3 | Steuben | IN | 11,700 | 0.34 | 34,594 | 1.02 | 3.60 |
| J4 | Lake | IN | 10,411 | 0.02 | 485,493 | 1.21 | 3.71 |
| J5 | Mariposa | CA | 10,000 | 0.58 | 17,203 | 0.28 | 3.08 |
| K1 | Portage | OH | 10,000 | 0.06 | 162,466 | 1.23 | 2.33 |
| K2 | Lake | IN | 8,978 | 0.02 | 485,493 | 1.26 | 3.29 |
| K3 | Kendall | IL | 8,100 | 0.06 | 128,990 | 0.58 | 3.63 |
| K4 | Park | MT | 8,000 | 0.48 | 16,606 | 0.73 | 4.36 |
| K5 | Monroe | FL | 6,413 | 0.09 | 74,228 | 0.65 | 2.00 |

Table S1. Catchment populations, county populations, mean flow (MGD), and mean sampling frequency for all sampling locations (samples per month). Sampling locations are also labeled according to their position in Figure 1.

| Figure 1 label | County | State | Spearman correlation (raw concentration) | Spearman correlation (normalized concentration) | Spearman correlation (three-sample average of normalized concentration) |
|---|---|---|---|---|---|
| A1 | Ramsey | MN | 0.81 | 0.57 | 0.68 |
| A2 | Sacramento | CA | 0.81 | 0.84 | 0.91 |

| | | | | | |
|---|---|---|---|---|---|
| A3 | Suffolk | MA | 0.79 | 0.86 | 0.89 |
| A4 | Suffolk | MA | 0.77 | 0.86 | 0.88 |
| A5 | Miami-Dade | FL | 0.86 | 0.82 | 0.84 |
| B1 | Miami-Dade | FL | 0.81 | 0.75 | 0.75 |
| B2 | Miami-Dade | FL | 0.82 | 0.78 | 0.76 |
| B3 | Multnomah | OR | 0.71 | 0.72 | 0.83 |
| B4 | New Castle | DE | 0.39 | 0.49 | 0.36 |
| B5 | Hamilton | TN | 0.49 | 0.69 | 0.69 |
| C1 | Shelby | TN | 0.57 | 0.87 | 0.93 |
| C2 | Arapahoe | CO | 0.89 | 0.80 | 0.86 |
| C3 | Union | NJ | 0.81 | 0.82 | 0.79 |
| C4 | Erie | PA | 0.88 | 0.92 | 0.93 |
| C5 | Lake | IN | 0.79 | 0.87 | 0.92 |
| D1 | Peoria | IL | 0.79 | 0.76 | 0.87 |
| D2 | Dauphin | PA | 0.84 | 0.76 | 0.77 |
| D3 | Stafford | VA | 0.80 | 0.68 | 0.76 |
| D4 | Jackson | OR | 0.46 | 0.55 | 0.71 |
| D5 | Wake | NC | 0.74 | 0.58 | 0.70 |
| E1 | Palm Beach | FL | 0.71 | 0.19 | 0.06 |
| E2 | Franklin | PA | 0.91 | 0.79 | 0.88 |
| E3 | Placer | CA | 0.79 | 0.75 | 0.76 |
| E4 | Wake | NC | 0.78 | 0.58 | 0.57 |
| E5 | Berkshire | MA | 0.84 | 0.84 | 0.81 |
| F1 | Middlesex | MA | 0.57 | 0.70 | 0.80 |
| F2 | Jackson | OR | 0.77 | 0.69 | 0.69 |
| F3 | Stafford | VA | 0.68 | 0.66 | 0.72 |
| F4 | Middlesex | MA | 0.53 | 0.74 | 0.77 |
| F5 | St. Lucie | FL | 0.59 | 0.67 | 0.62 |
| G1 | Fairfield | CT | 0.64 | 0.77 | 0.82 |
| G2 | Nassau | NY | 0.86 | 0.86 | 0.87 |
| G3 | Essex | MA | 0.85 | 0.89 | 0.92 |
| G4 | Hampshire | MA | 0.54 | 0.43 | 0.41 |
| G5 | Jackson | OR | 0.59 | 0.64 | 0.72 |
| H1 | Middlesex | MA | 0.33 | 0.59 | 0.75 |
| H2 | Indiana | PA | 0.78 | 0.84 | 0.89 |
| H3 | Lake | IN | 0.71 | 0.68 | 0.70 |

| | | | | | |
|---|---|---|---|---|---|
| H4 | Lake | IN | 0.66 | 0.66 | 0.84 |
| H5 | Elko | NV | 0.79 | 0.79 | 0.89 |
| I1 | Umatilla | OR | 0.75 | 0.85 | 0.88 |
| I2 | Nantucket | MA | 0.84 | 0.85 | 0.85 |
| I3 | Hampshire | MA | 0.73 | 0.79 | 0.94 |
| I4 | Portage | OH | 0.81 | 0.84 | 0.93 |
| I5 | Monterey | CA | 0.54 | 0.56 | 0.58 |
| J1 | Montgomery | PA | 0.78 | 0.71 | 0.66 |
| J2 | Middlesex | MA | 0.48 | 0.56 | 0.58 |
| J3 | Steuben | IN | 0.66 | 0.63 | 0.59 |
| J4 | Lake | IN | 0.77 | 0.71 | 0.71 |
| J5 | Mariposa | CA | 0.56 | 0.61 | 0.49 |
| K1 | Portage | OH | 0.89 | 0.89 | 0.89 |
| K2 | Lake | IN | 0.81 | 0.78 | 0.63 |
| K3 | Kendall | IL | 0.30 | 0.31 | 0.44 |
| K4 | Park | MT | 0.82 | 0.81 | 0.86 |
| K5 | Monroe | FL | 0.86 | 0.74 | 0.68 |

Table S2. Correlations calculated between wastewater concentrations and reported clinical cases for all sampling locations.

| Month | Spearman correlation (normalized concentration) | Spearman correlation (raw concentration) | Number of states |
|---|---|---|---|
| April 2020 | 0.58 | 0.50 | 16 |
| May 2020 | 0.42 | 0.55 | 19 |
| June 2020 | 0.79 | 0.62 | 19 |
| July 2020 | 0.90 | 0.77 | 18 |
| August 2020 | 0.86 | 0.80 | 19 |
| September 2020 | 0.51 | 0.35 | 19 |
| October 2020 | 0.62 | 0.47 | 19 |
| November 2020 | 0.82 | 0.72 | 19 |
| December 2020 | 0.56 | 0.20 | 19 |
| January 2021 | 0.59 | 0.37 | 16 |
| February 2021 | 0.51 | 0.56 | 14 |
| March 2021 | 0.79 | 0.57 | 14 |
| April 2021 | 0.26 | 0.25 | 13 |

| | | | |
|---|---|---|---|
| May 2021 | 0.82 | 0.65 | 13 |

Table S3. Correlations between wastewater concentrations and clinical cases across states. Wastewater concentrations were averaged across all locations per state each month. Clinical cases were averaged within each county with at least one respective sampling location and then also averaged per state each month. Correlations were calculated with the raw and normalized wastewater concentrations. “Number of states” indicates the number of states which had a least one sampling location for the respective month (i.e. which contributed to that month’s correlation calculation).

### Supplementary File 2. Detailed time series for all profiled locations.

**Supp_File_2 - time_series_all_with_details.pdf**

Each plot shows one sampling location’s time series. (Left axis) Blue line: centered 3-sample average of normalized wastewater concentrations (genome copies/L); blue dots: individual normalized wastewater concentration measurements (genome copies/L). (Right axis) Orange line: centered 7-day average of daily new cases in the respective county (new cases); gray bars: reported new daily cases (new cases). Y-axes are normalized to the maximum of each time series. Plots are titled with their location in Figure 1 and their primary county.
