## Supplemental File 2 for "Nationwide trends in COVID-19 cases and SARS-CoV-2 wastewater concentrations in the United States"

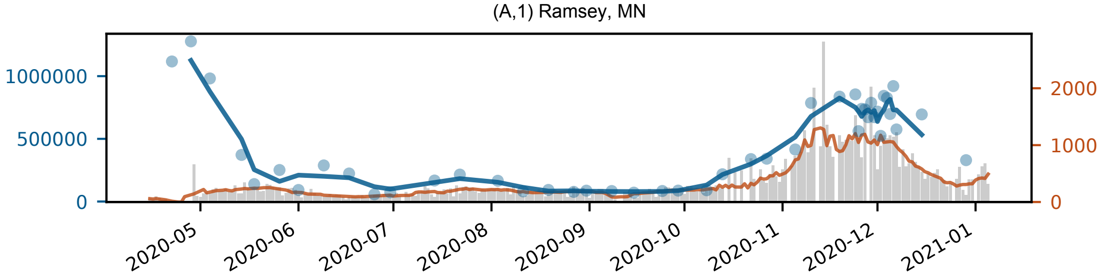
(A,1) Ramsey, MN
1000000
500000
0
2000
1000
0
2020-05
2020-06
2020-07
2020-08
2020-09
2020-10
2020-11
2020-12
2021-01

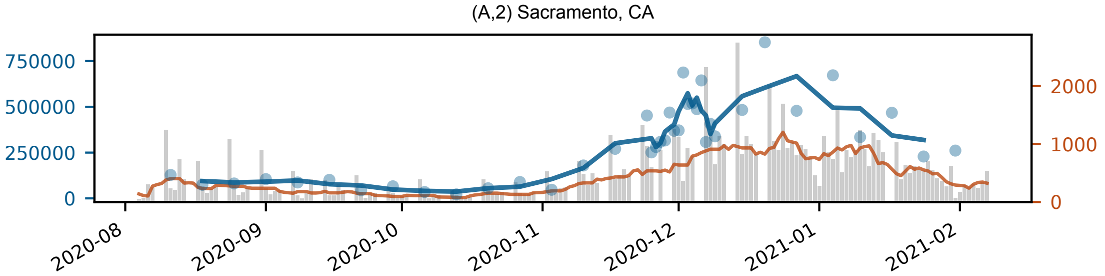
(A,2) Sacramento, CA
750000
500000
250000
0
2000
1000
0
2020-08
2020-09
2020-10
2020-11
2020-12
2021-01
2021-02

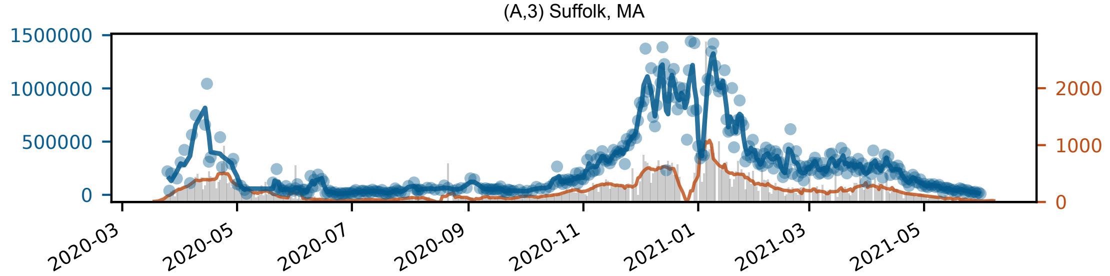

(A,3) Suffolk, MA
1500000
1000000
500000
0
2000
1000
0
2020-03
2020-05
2020-07
2020-09
2020-11
2021-01
2021-03
2021-05

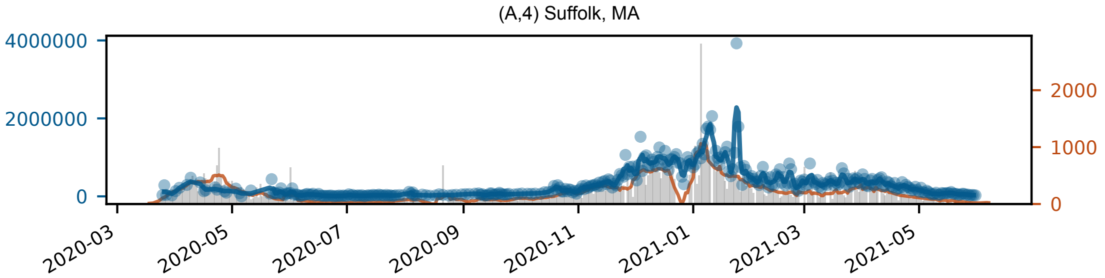

(A,4) Suffolk, MA
4000000
2000000
0
2000
1000
0
2020-03
2020-05
2020-07
2020-09
2020-11
2021-01
2021-03
2021-05

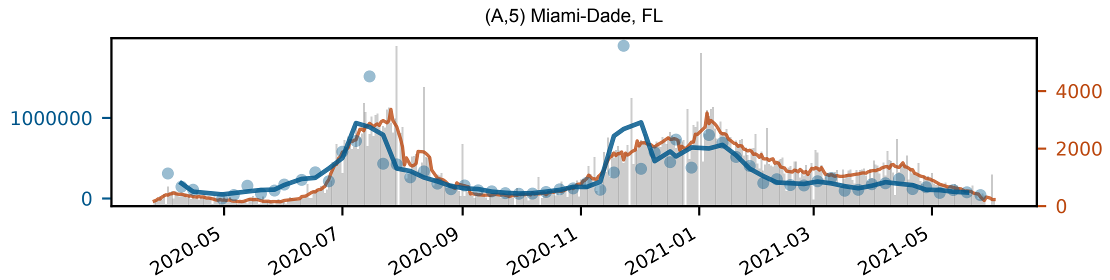
(A,5) Miami-Dade, FL
1000000
0
4000
2000
0
2020-05
2020-07
2020-09
2020-11
2021-01
2021-03
2021-05

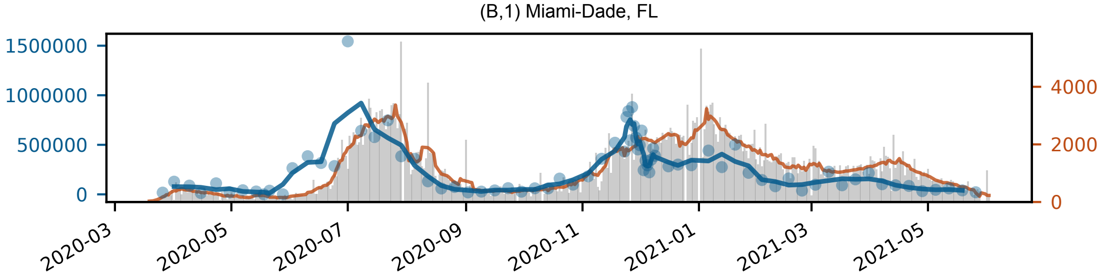

(B,1) Miami-Dade, FL
1500000
1000000
500000
0
4000
2000
0
2020-03
2020-05
2020-07
2020-09
2020-11
2021-01
2021-03
2021-05

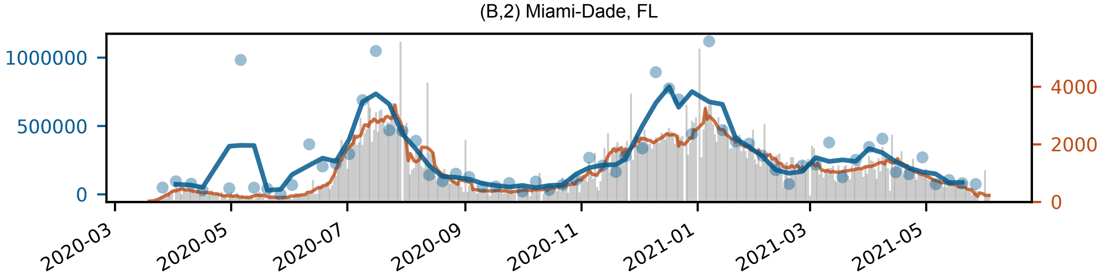

(B,2) Miami-Dade, FL
1000000
500000
0
4000
2000
0
2020-03
2020-05
2020-07
2020-09
2020-11
2021-01
2021-03
2021-05

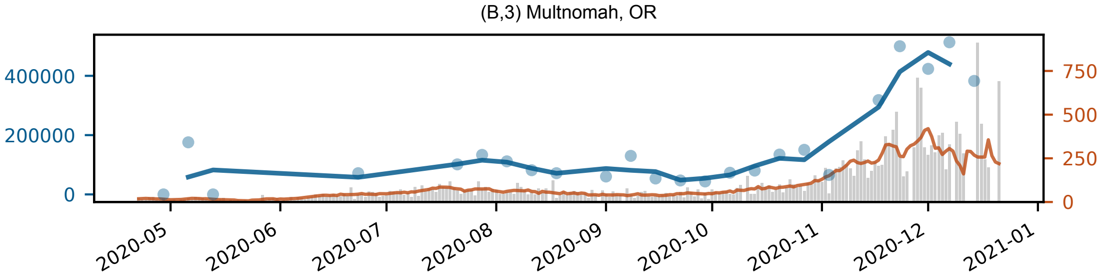

(B,3) Multnomah, OR
400000
200000
0
750
500
250
0
2020-05
2020-06
2020-07
2020-08
2020-09
2020-10
2020-11
2020-12
2021-01

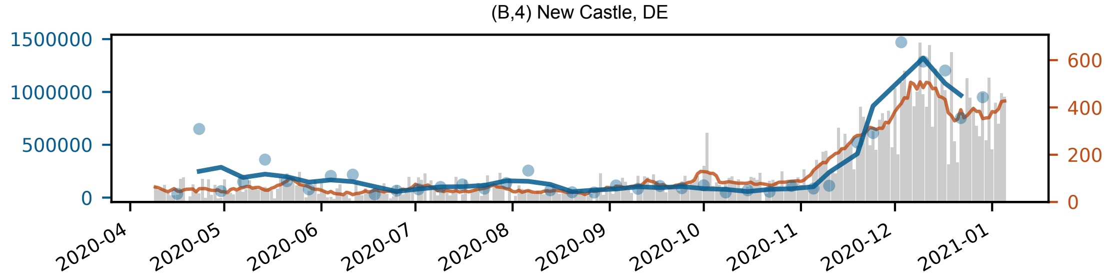

(B,4) New Castle, DE
1500000
1000000
500000
0
600
400
200
0
2020-04
2020-05
2020-06
2020-07
2020-08
2020-09
2020-10
2020-11
2020-12
2021-01

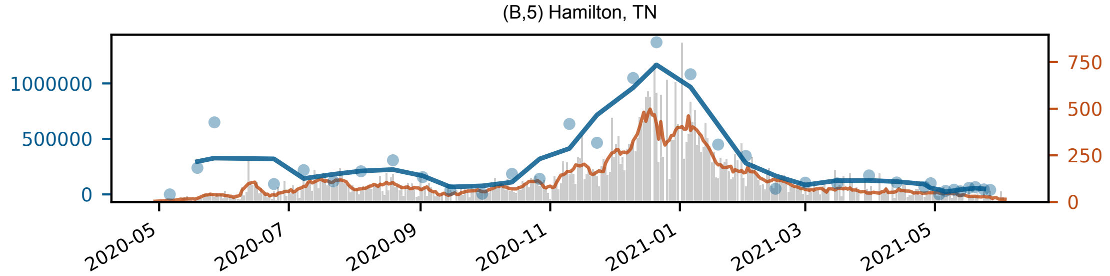

(B,5) Hamilton, TN
1000000
500000
0
750
500
250
0
2020-05
2020-07
2020-09
2020-11
2021-01
2021-03
2021-05

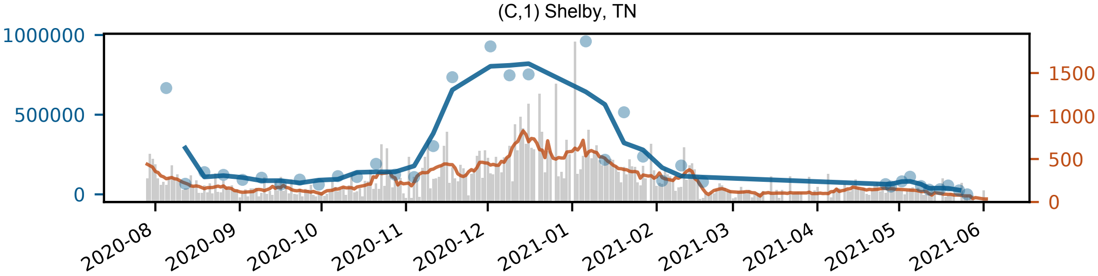

(C,1) Shelby, TN
1000000
500000
0
1500
1000
500
0
2020-08
2020-09
2020-10
2020-11
2020-12
2021-01
2021-02
2021-03
2021-04
2021-05
2021-06

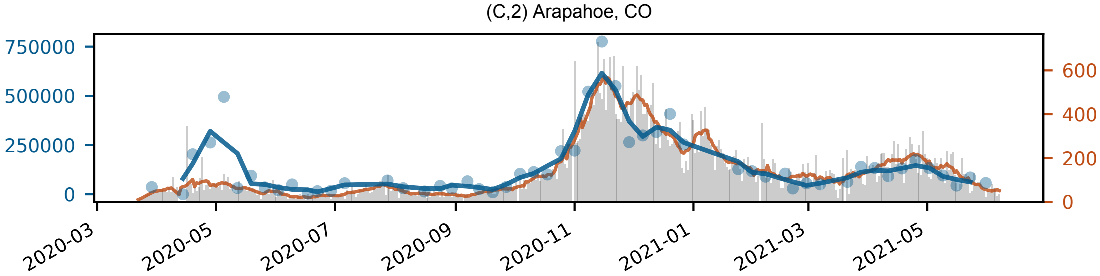

(C,2) Arapahoe, CO
750000
500000
250000
0
600
400
200
0
2020-03
2020-05
2020-07
2020-09
2020-11
2021-01
2021-03
2021-05

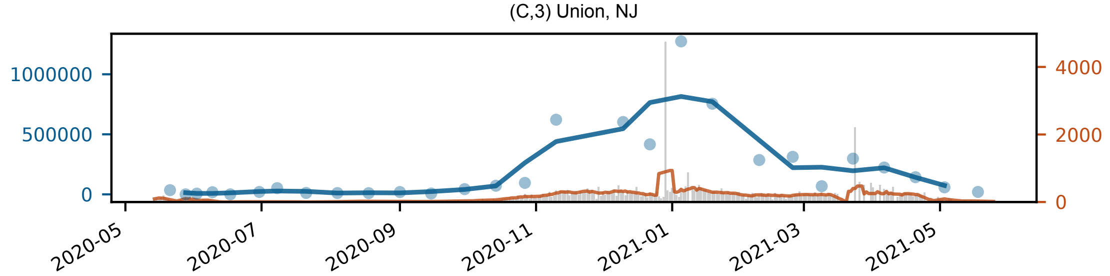

(C,3) Union, NJ
1000000
500000
0
4000
2000
0
2020-05
2020-07
2020-09
2020-11
2021-01
2021-03
2021-05

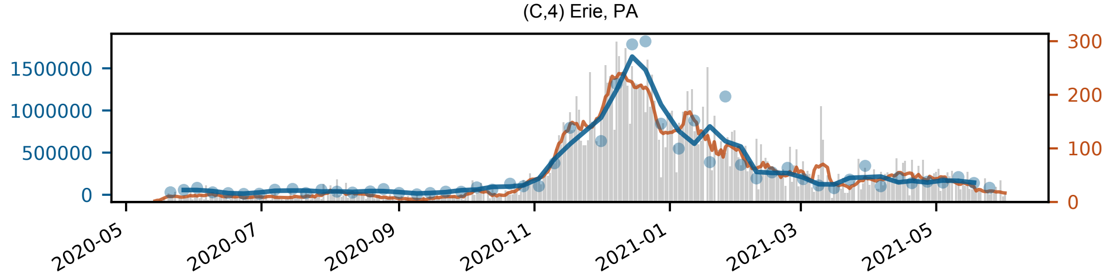

(C,4) Erie, PA
1500000
1000000
500000
0
300
200
100
0
2020-05
2020-07
2020-09
2020-11
2021-01
2021-03
2021-05

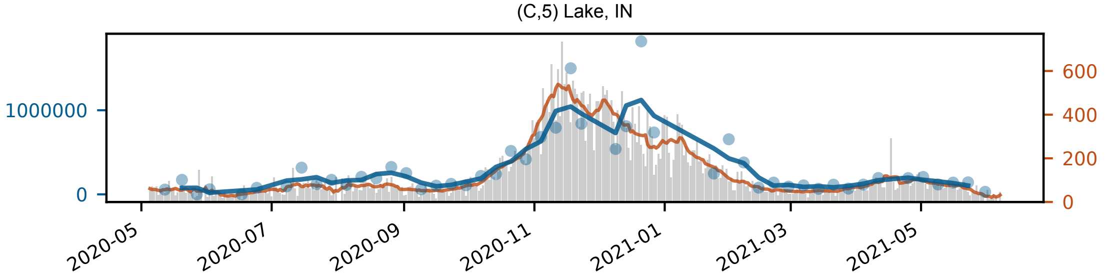

(C,5) Lake, IN
1000000
0
600
400
200
0
2020-05
2020-07
2020-09
2020-11
2021-01
2021-03
2021-05

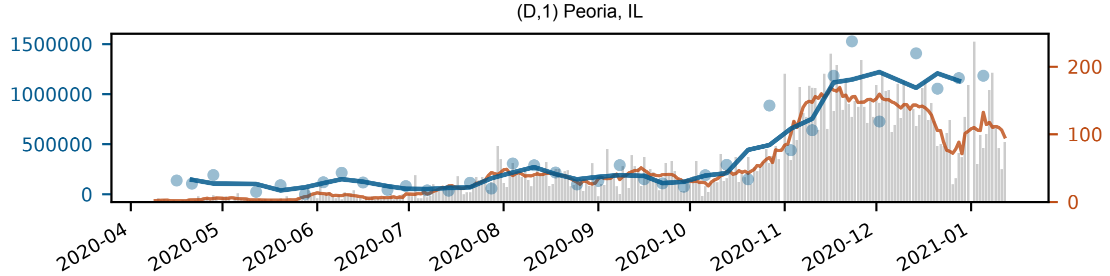

(D,1) Peoria, IL
1500000
1000000
500000
0
200
100
0
2020-04
2020-05
2020-06
2020-07
2020-08
2020-09
2020-10
2020-11
2020-12
2021-01

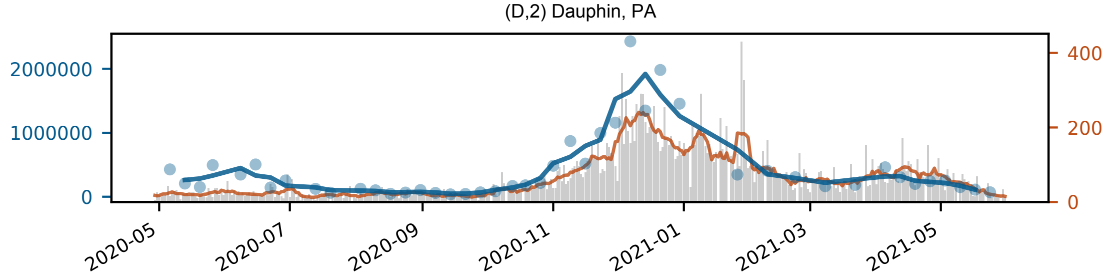

(D,2) Dauphin, PA
2000000
1000000
0
400
200
0
2020-05
2020-07
2020-09
2020-11
2021-01
2021-03
2021-05

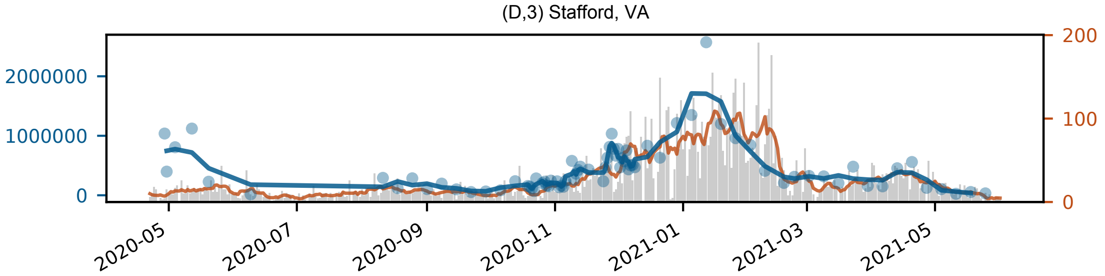

(D,3) Stafford, VA
2000000
1000000
0
200
100
0
2020-05
2020-07
2020-09
2020-11
2021-01
2021-03
2021-05

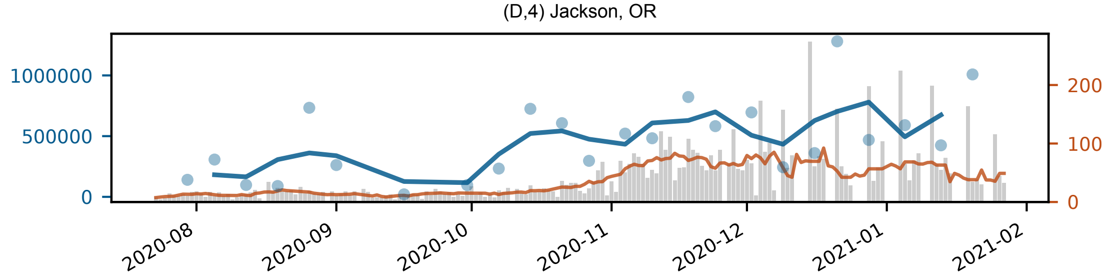

(D,4) Jackson, OR
1000000
500000
0
200
100
0
2020-08
2020-09
2020-10
2020-11
2020-12
2021-01
2021-02

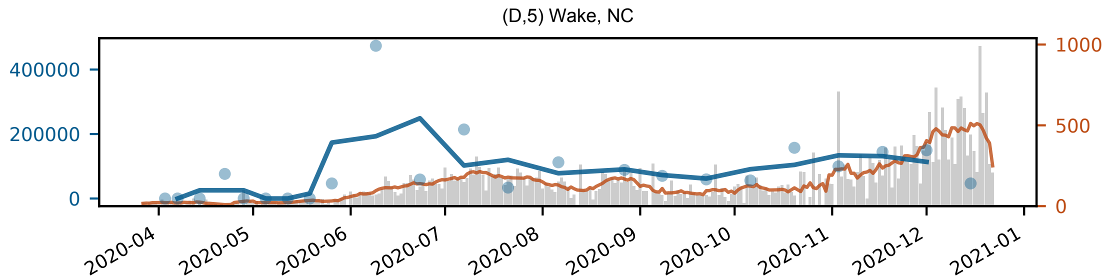

(D,5) Wake, NC
400000
200000
0
1000
500
0
2020-04
2020-05
2020-06
2020-07
2020-08
2020-09
2020-10
2020-11
2020-12
2021-01

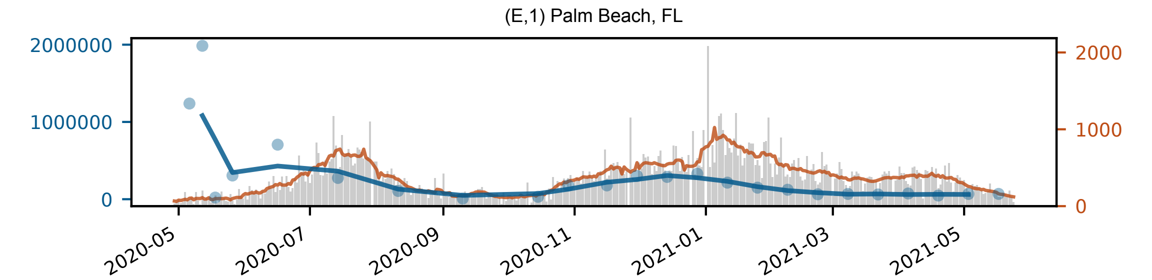

(E,1) Palm Beach, FL
2000000
1000000
0
2000
1000
0
2020-05
2020-07
2020-09
2020-11
2021-01
2021-03
2021-05

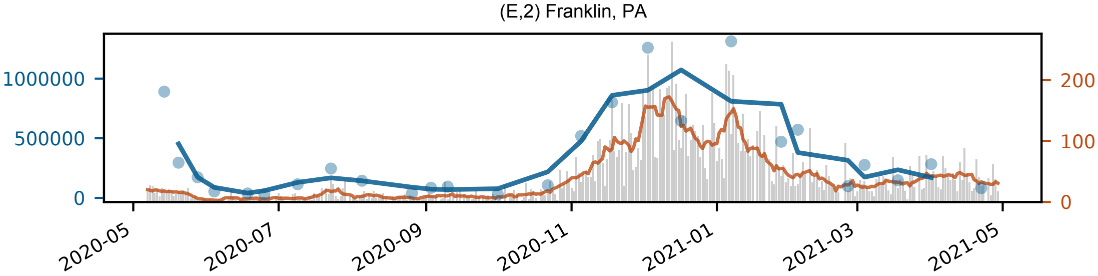

(E,2) Franklin, PA
1000000
500000
0
200
100
0
2020-05
2020-07
2020-09
2020-11
2021-01
2021-03
2021-05

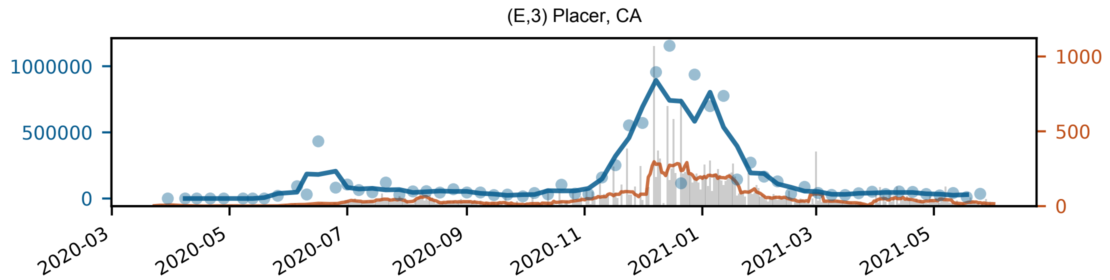
(E,3) Placer, CA
1000000
500000
0
1000
500
0
2020-03
2020-05
2020-07
2020-09
2020-11
2021-01
2021-03
2021-05

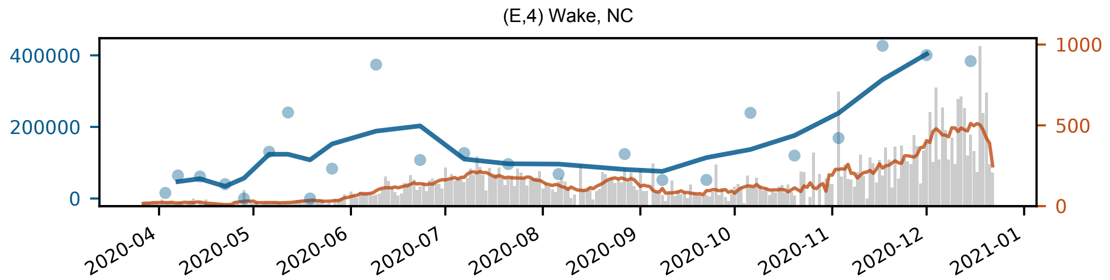

(E,4) Wake, NC
400000
200000
0
1000
500
0
2020-04
2020-05
2020-06
2020-07
2020-08
2020-09
2020-10
2020-11
2020-12
2021-01

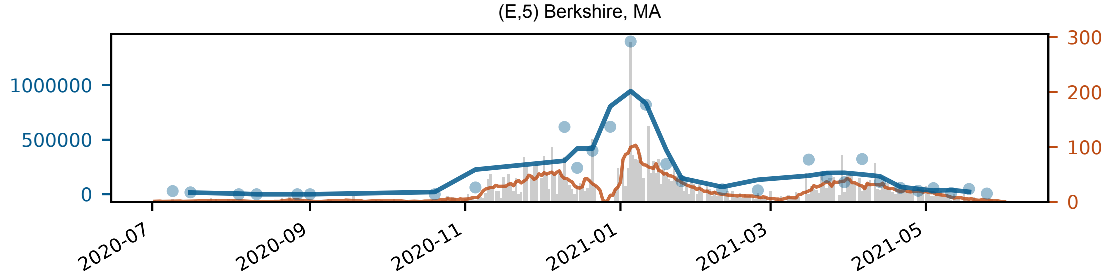

(E,5) Berkshire, MA
1000000
500000
0
300
200
100
0
2020-07
2020-09
2020-11
2021-01
2021-03
2021-05

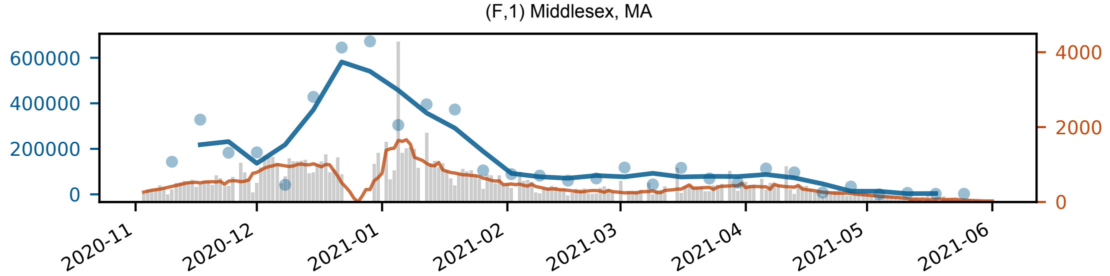

(F,1) Middlesex, MA
600000
400000
200000
0
4000
2000
0
2020-11
2020-12
2021-01
2021-02
2021-03
2021-04
2021-05
2021-06

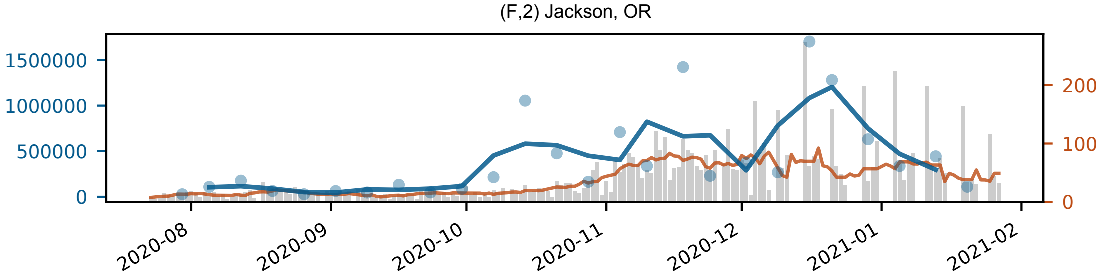

(F,2) Jackson, OR
1500000
1000000
500000
0
200
100
0
2020-08
2020-09
2020-10
2020-11
2020-12
2021-01
2021-02

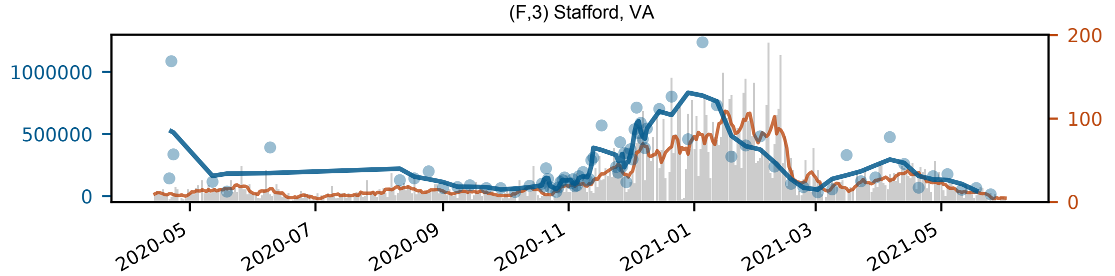

(F,3) Stafford, VA
1000000
500000
0
200
100
0
2020-05
2020-07
2020-09
2020-11
2021-01
2021-03
2021-05

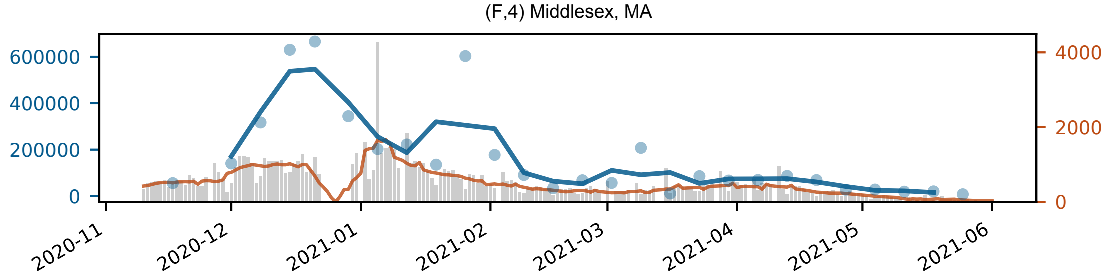

(F,4) Middlesex, MA
600000
400000
200000
0
4000
2000
0
2020-11
2020-12
2021-01
2021-02
2021-03
2021-04
2021-05
2021-06

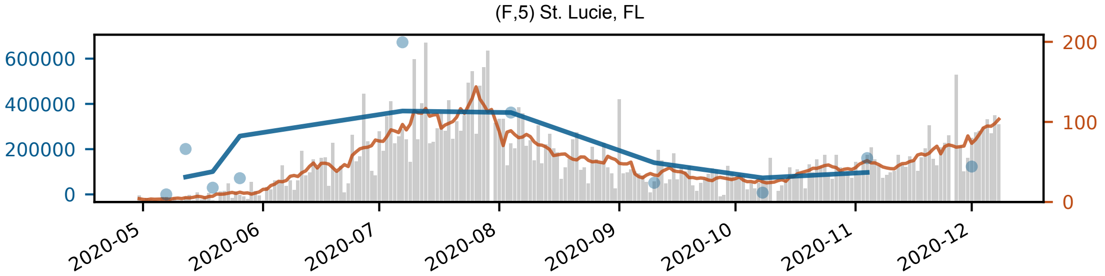

(F,5) St. Lucie, FL
600000
400000
200000
0
200
100
0
2020-05
2020-06
2020-07
2020-08
2020-09
2020-10
2020-11
2020-12

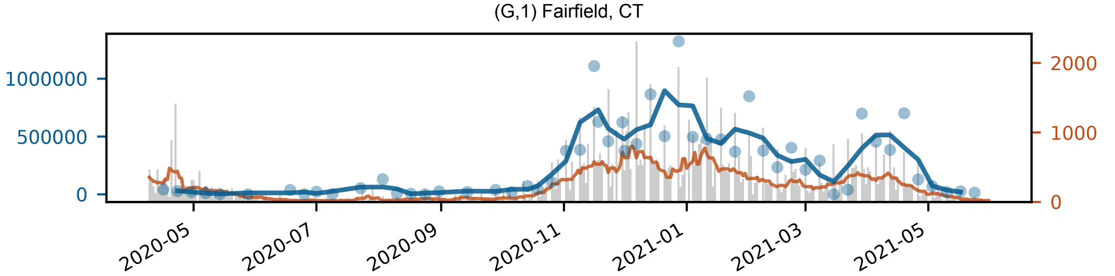
(G,1) Fairfield, CT
1000000
500000
0
2000
1000
0
2020-05
2020-07
2020-09
2020-11
2021-01
2021-03
2021-05

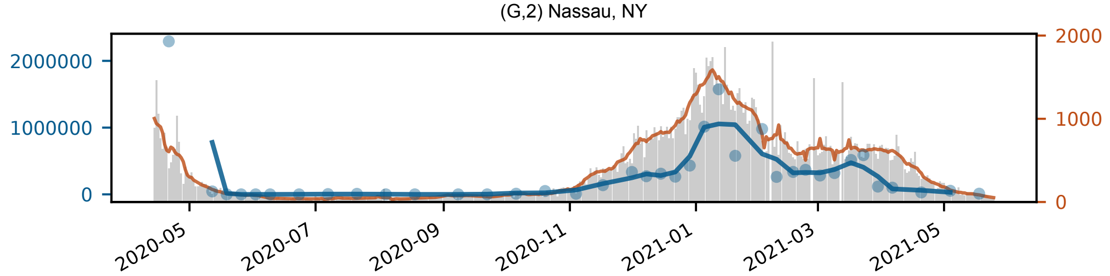

(G,2) Nassau, NY
2000000
1000000
0
2000
1000
0
2020-05
2020-07
2020-09
2020-11
2021-01
2021-03
2021-05

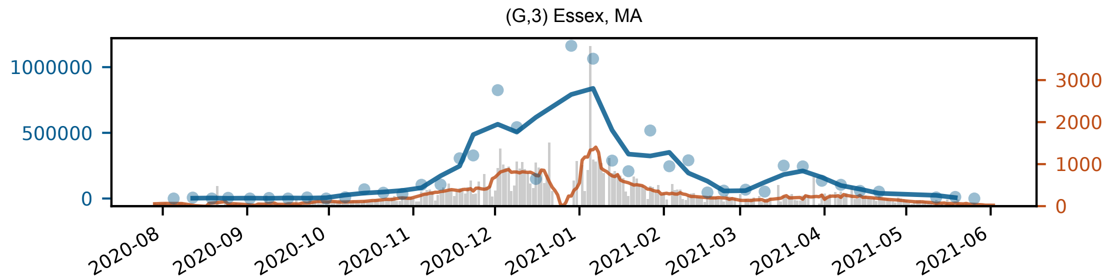

(G,3) Essex, MA
1000000
500000
0
3000
2000
1000
0
2020-08
2020-09
2020-10
2020-11
2020-12
2021-01
2021-02
2021-03
2021-04
2021-05
2021-06

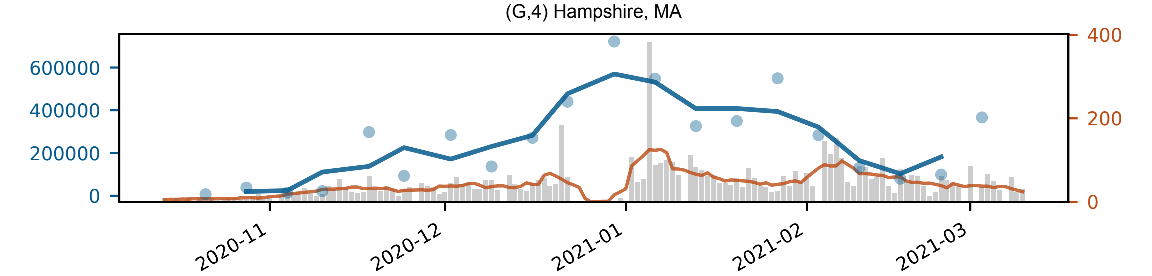

(G,4) Hampshire, MA
600000
400000
200000
0
400
200
0
2020-11
2020-12
2021-01
2021-02
2021-03

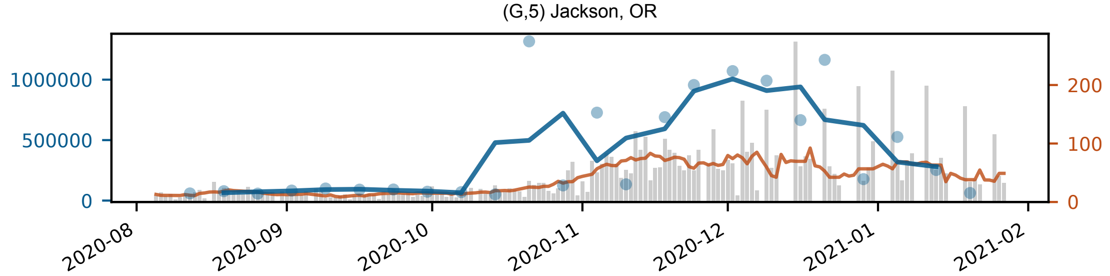

(G,5) Jackson, OR
1000000
500000
0
200
100
0
2020-08
2020-09
2020-10
2020-11
2020-12
2021-01
2021-02

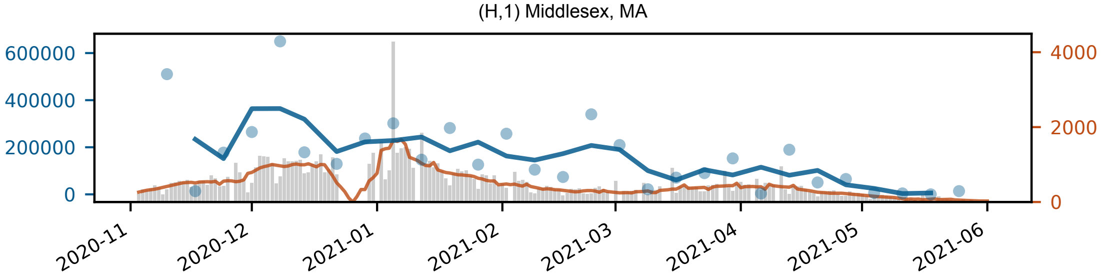

(H,1) Middlesex, MA
600000
400000
200000
0
4000
2000
0
2020-11
2020-12
2021-01
2021-02
2021-03
2021-04
2021-05
2021-06

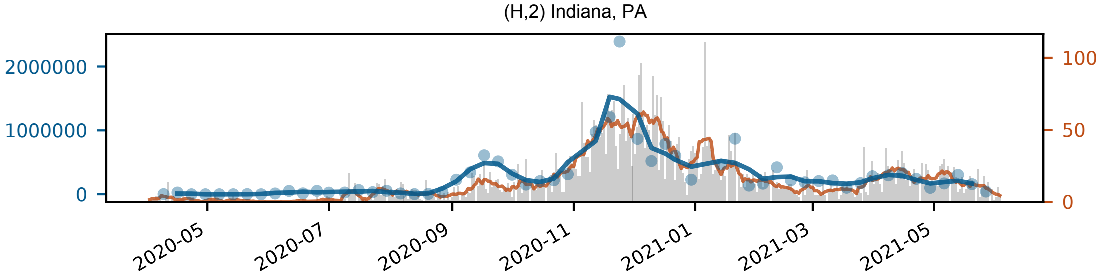

(H,2) Indiana, PA
2000000
1000000
0
100
50
0
2020-05
2020-07
2020-09
2020-11
2021-01
2021-03
2021-05

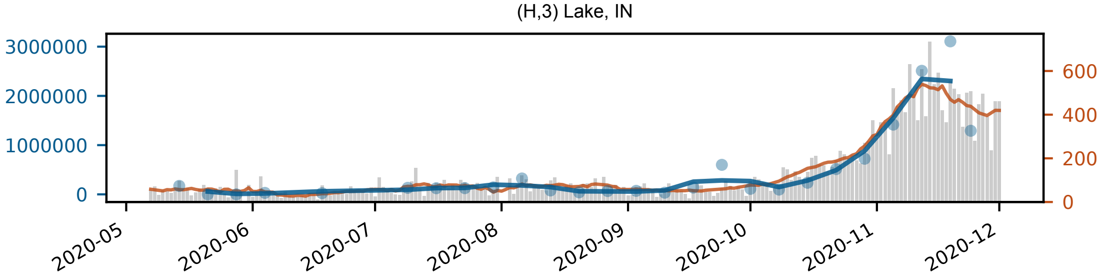

(H,3) Lake, IN
3000000
2000000
1000000
0
600
400
200
0
2020-05
2020-06
2020-07
2020-08
2020-09
2020-10
2020-11
2020-12

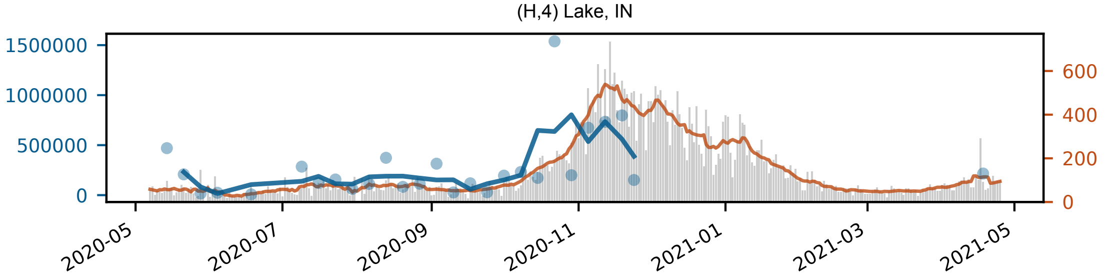

(H,4) Lake, IN
1500000
1000000
500000
0
600
400
200
0
2020-05
2020-07
2020-09
2020-11
2021-01
2021-03
2021-05

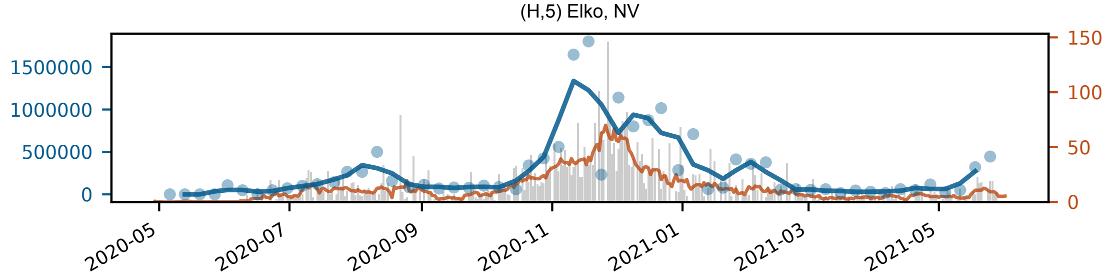

(H,5) Elko, NV
1500000
1000000
500000
0
150
100
50
0
2020-05
2020-07
2020-09
2020-11
2021-01
2021-03
2021-05

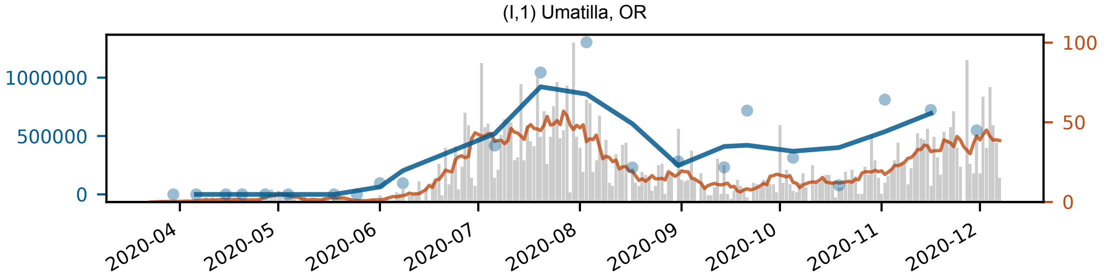
(I,1) Umatilla, OR
1000000
500000
0
100
50
0
2020-04
2020-05
2020-06
2020-07
2020-08
2020-09
2020-10
2020-11
2020-12

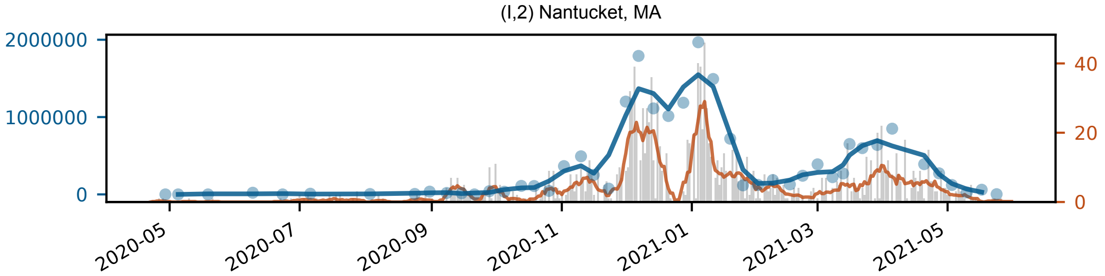

(I,2) Nantucket, MA
2000000
1000000
0
40
20
0
2020-05
2020-07
2020-09
2020-11
2021-01
2021-03
2021-05

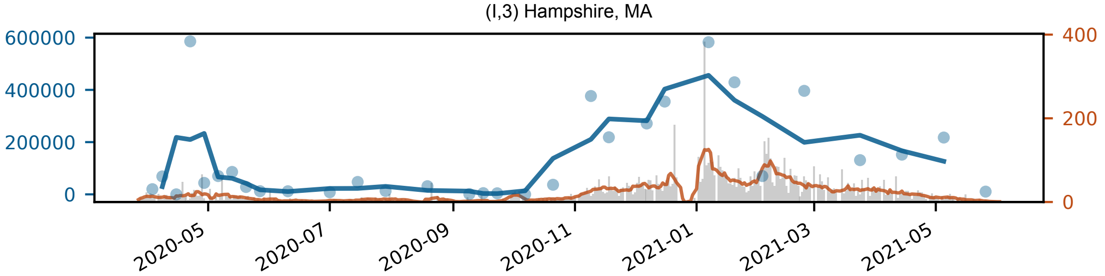
(I,3) Hampshire, MA
600000
400000
200000
0
400
200
0
2020-05
2020-07
2020-09
2020-11
2021-01
2021-03
2021-05

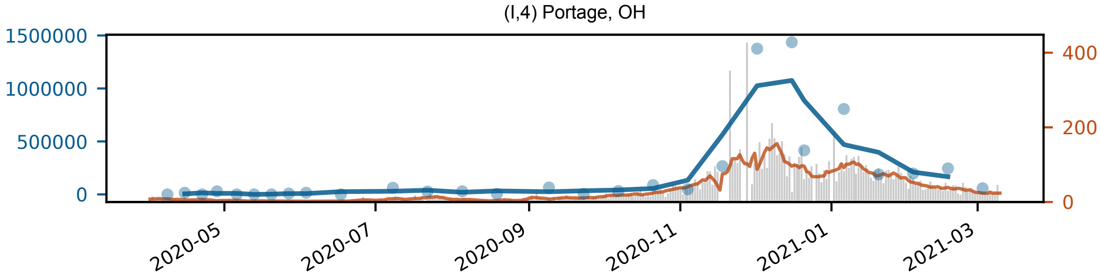
(I,4) Portage, OH
1500000
1000000
500000
0
400
200
0
2020-05
2020-07
2020-09
2020-11
2021-01
2021-03

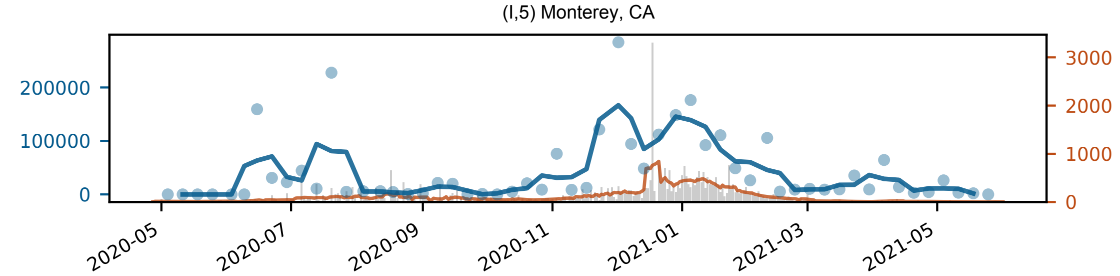

(I,5) Monterey, CA
200000
100000
0
3000
2000
1000
0
2020-05
2020-07
2020-09
2020-11
2021-01
2021-03
2021-05

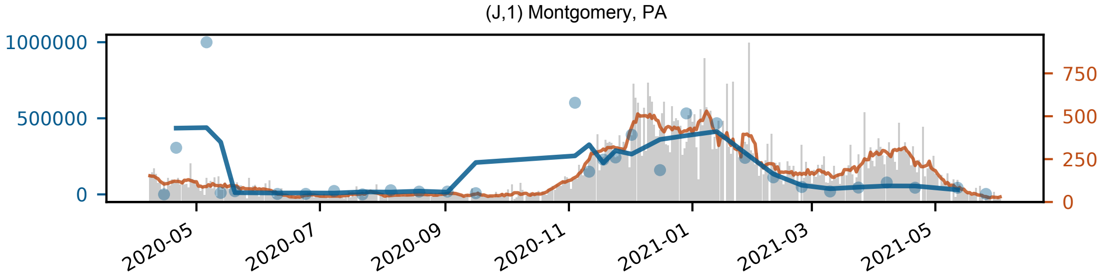

(J,1) Montgomery, PA
1000000
500000
0
750
500
250
0
2020-05
2020-07
2020-09
2020-11
2021-01
2021-03
2021-05

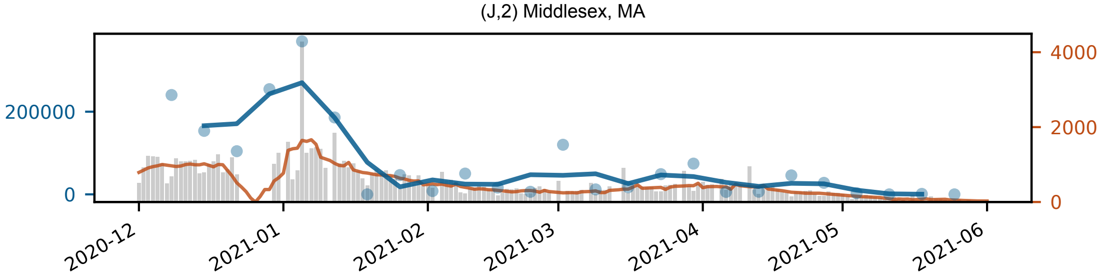

(J,2) Middlesex, MA
200000
0
4000
2000
0
2020-12
2021-01
2021-02
2021-03
2021-04
2021-05
2021-06

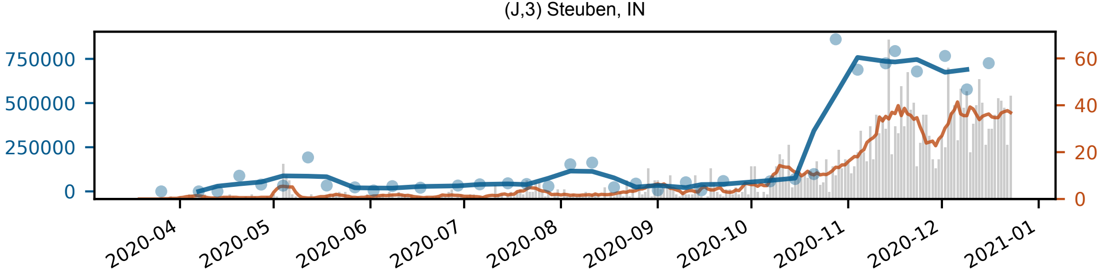

(J,3) Steuben, IN
750000
500000
250000
0
60
40
20
0
2020-04
2020-05
2020-06
2020-07
2020-08
2020-09
2020-10
2020-11
2020-12
2021-01

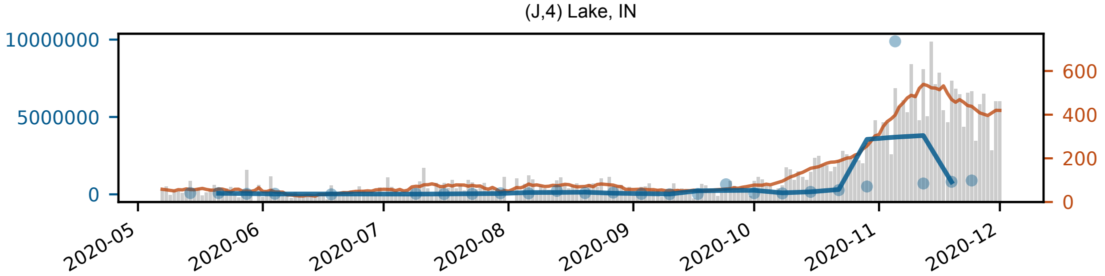

(J,4) Lake, IN
10000000
5000000
0
600
400
200
0
2020-05
2020-06
2020-07
2020-08
2020-09
2020-10
2020-11
2020-12

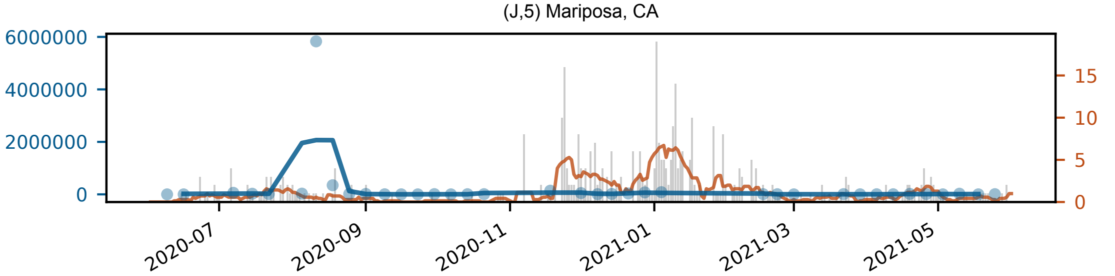
(J,5) Mariposa, CA
6000000
4000000
2000000
0
15
10
5
0
2020-07
2020-09
2020-11
2021-01
2021-03
2021-05

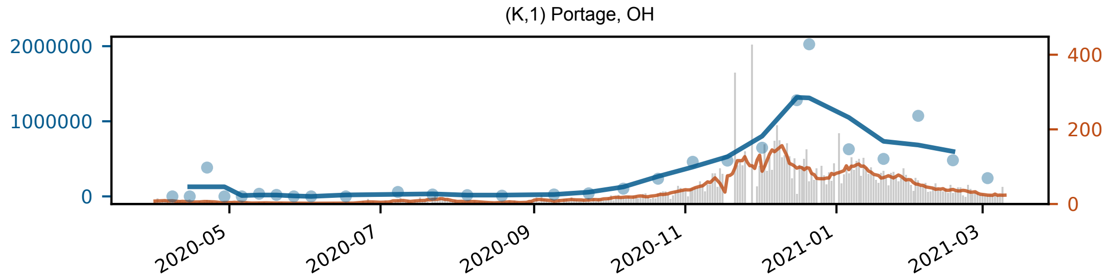

(K,1) Portage, OH
2000000
1000000
0
400
200
0
2020-05
2020-07
2020-09
2020-11
2021-01
2021-03

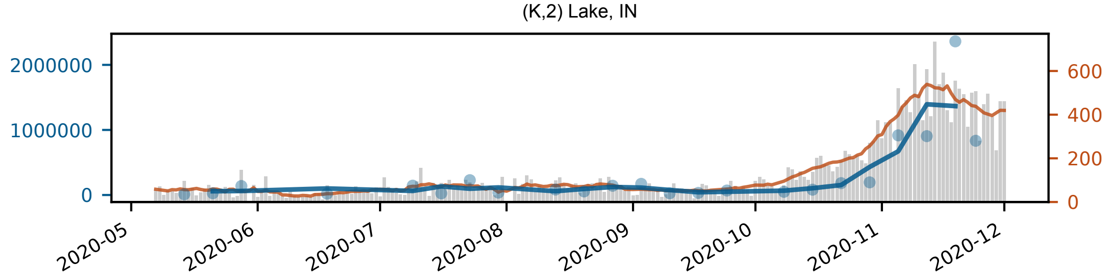

(K,2) Lake, IN
2000000
1000000
0
600
400
200
0
2020-05
2020-06
2020-07
2020-08
2020-09
2020-10
2020-11
2020-12

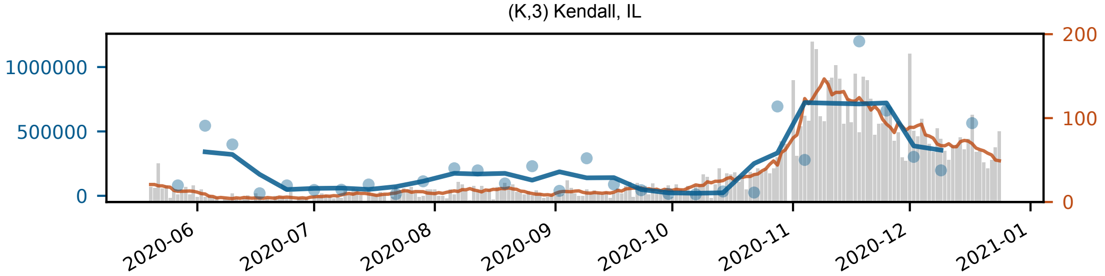

(K,3) Kendall, IL
1000000
500000
0
200
100
0
2020-06
2020-07
2020-08
2020-09
2020-10
2020-11
2020-12
2021-01

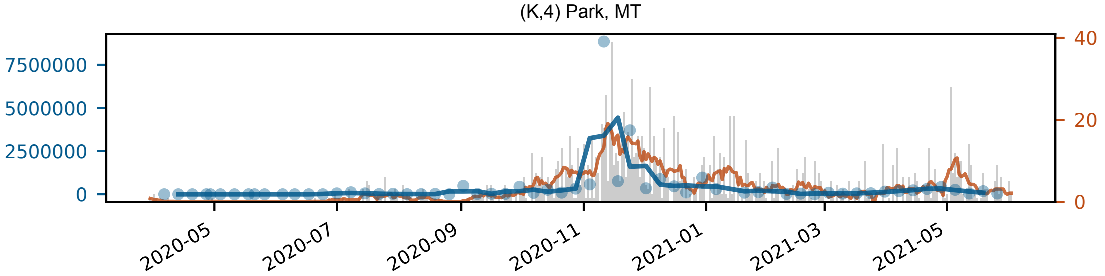

(K,4) Park, MT
7500000
5000000
2500000
0
40
20
0
2020-05
2020-07
2020-09
2020-11
2021-01
2021-03
2021-05

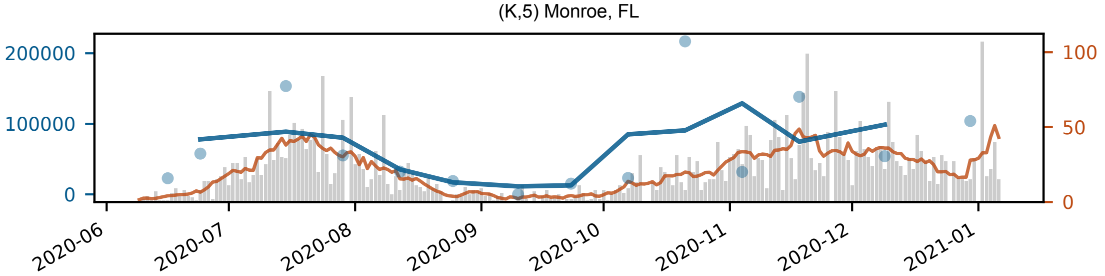

(K,5) Monroe, FL
200000
100000
0
100
50
0
2020-06
2020-07
2020-08
2020-09
2020-10
2020-11
2020-12
2021-01
